# Accounting for sex differences variability in the design of sex-adapted cancer treatments

**DOI:** 10.1101/2023.04.22.23288966

**Authors:** Wei Yang, Joshua B Rubin

## Abstract

The significant sex differences that exist in cancer mechanisms, incidence, and survival, have yet to impact clinical practice. We hypothesized that one barrier to translation is that sex differences in cancer phenotypes resemble sex differences in height: highly overlapping, but distinct, male and female population distributions that vary continuously between female- and male-biased extremes. A consequence of this variance is that sex-specific treatments are rendered unrealistic, and our translational goal should be adaptation of treatment to the unique mix of sex-biased mechanisms that are present in each patient. To develop a tool that could advance this goal, we applied a Bayesian Nearest Neighbor (BNN) analysis to 8370 cancer transcriptomes from 26 different adult and 4 different pediatric cancer types to establish patient-specific Transcriptomic Sex Indices (TSI). TSI precisely partitions an individual patient’s whole transcriptome into female- and male-biased components such that cancer type, patient sex, and transcriptomics, provide a novel and patient-specific mechanistic identifier that can be used for sex-adapted, precision cancer treatment planning.

## Introduction

Significant sex differences in cancer incidence and mortality are recognized to be the norm with the majority of cancers exhibiting male to female incidence ratios ranging from 1.26:1 to 4.86:1 [1]. Recent analysis of over 14 million cases from the Cancer Registry, representing 99.9% of the cancer population of the United States confirmed an overall predominance of male cancer cases [2]. An accompanying analysis of survival data from 3.7 million cases in the SEER database, representing approximately 28% of the cancer population, confirmed that mortality rates are higher for males compared to females [2]. These clinically important sex differences are concordant with described sex differences in cell biology including, response to genotoxic stress [3, 4], DNA repair [5, 6], mutational burden [7, 8], metabolism [9, 10], and cell cycle regulation [11-13], as well as in systems biology including: immunity [14], metabolism [15, 16], tissue repair [17, 18], and longevity [19, 20]. This suggests that therapies for all cancer patients may be advanced by a realistic translation of sex differences into clinical practice.

An obstacle to this end is our incomplete understanding of how sex and its genetic, epigenetic, and hormonal foundations, mechanistically interact with each of the hallmark pathways of cancer to influence its genesis, progression, and response to treatment. A second major obstacle lies in the nature of biological sex differences. Most sex differences are not dichotomous or sexually dimorphic, like ovaries versus testes. Instead, most sex differences are more akin to height, a complex trait that varies continuously between the shortest females and the tallest males, but is intermediate for most people [21]. Thus, investigating the influence of sex on cancer and applying that knowledge in the clinical setting is complicated by the varying interactions between chromosomal and gonadal sex, and multiple other cellular, tissue, and systemic features that comprise an individual’s cancer phenotype.

While many males and females possess intermediate heights between the female (shortest) and male (tallest) poles, the mechanisms that promote intermediate heights may differ between the sexes.

Height-determining mechanisms underlying a female height that is taller than their extreme or pole, are likely to be different from those in males that are shorter than their pole. Thus, targeting even similar cancer phenotypes in male and female patients they may benefit from sex-adapted treatment plans.

We sought a method by which to measure sex-biased mechanisms underlying cancer phenotypes. To do this, we used a Bayesian Nearest Neighbor (BNN) analysis to identify the poles of male and female transcriptional phenotypes in pediatric and adult cancers. We calculated a patient-specific Transcriptomic Sex Index (TSI) value based on the BNN analysis that precisely located each individual between the female (smallest TSI values) and male (highest TSI values) poles. With this analysis, we determined that most cancer diagnostic groups exhibit sex-biased transcriptomes in which cell cycle regulation and immunity/inflammation are the pathways most commonly associated with the male and female poles, respectively. In addition, we identified differing mechanisms associated with midrange TSI values in female and male patients. We conclude that even when males and females share a phenotypic feature, the mechanisms underlying that phenotype can be sex-biased. This is consistent with published analyses demonstrating that even when genes are equally expressed in males and females, they can exhibit different correlations to cancer mechanisms, treatment responses, and survival [22, 23]. We expect that the TSI approach will advance sex differences research and provide a paradigm for using an individual’s entire transcriptome for planning their individualized cancer treatment.

## Methods

The use of publicly accessible human datasets for research has been approved by the Washington University Institutional Review Board (IRB# 201102299).

### Inferring Transcriptomic Sex Index (TSI) Using Bayesian Nearest Neighbors

To identify sex-biased gene expression patterns in cancer, we first downloaded the TCGA pan-cancer transcriptome data (gene expression RNAseq - Batch effects normalized mRNA data) from https://pancanatlas.xenahubs.net, the Kids First neuroblastoma data from dbGaP (https://www.ncbi.nlm.nih.gov/projects/gap/cgi-bin/study.cgi?study_id=phs001436.v1.p1) and the Children’s Brain Tumor Network brain tumor data from https://cbtn.org. Sample sizes and number of genes are listed in **Table 1**. We excluded non-malignancies, cancer types with highly skewed numbers of male or female cases, and those cancers with low incidence (<45 cases in the datasets). The remaining 26 adult and four pediatric cancers have sample sizes ranging from 45 to 572, with male samples comprising 27.2% to 84.2% of each cancer type. The total cases examined were 8370 (4927 Males, 3443 (58.9%) Females (41.1%)).

**Table 1:** Case Data

| **PANCAN Types** | **genes** | **Male** | **Female** | **Total** | **% Male** |
| --- | --- | --- | --- | --- | --- |
| THCA | 20531 | 157 | 415 | 572 | 27.4 |
| ACC | 20531 | 31 | 48 | 79 | 39.2 |
| PCPG | 20531 | 84 | 103 | 187 | 44.9 |
| SARC | 20531 | 120 | 145 | 265 | 45.3 |
| DLBC | 20531 | 22 | 26 | 48 | 45.8 |
| LUAD | 20531 | 265 | 311 | 576 | 46.0 |
| CHOL | 20531 | 22 | 23 | 45 | 48.9 |
| THYM | 20531 | 63 | 59 | 122 | 51.6 |
| COAD | 17507 | 257 | 235 | 492 | 52.2 |
| READ | 17507 | 90 | 80 | 170 | 52.9 |
| LAML | 16765 | 93 | 80 | 173 | 53.8 |
| LGG | 20531 | 291 | 238 | 529 | 55.0 |
| PAAD | 20531 | 101 | 82 | 183 | 55.2 |
| UVM | 20531 | 45 | 35 | 80 | 56.3 |
| KICH | 20531 | 52 | 39 | 91 | 57.1 |
| <u>GBM525</u> | 8720 | 320 | 205 | 525 | 61.0 |
| SKCM | 20531 | 293 | 180 | 473 | 61.9 |
| GBM | 20531 | 107 | 59 | 166 | 64.5 |
| STAD | 16765 | 291 | 159 | 450 | 64.7 |
| KIRC | 20531 | 398 | 208 | 606 | 65.7 |
| LIHC | 20531 | 280 | 143 | 423 | 66.2 |
| BLCA | 20531 | 311 | 116 | 427 | 72.8 |
| KIRP | 20531 | 236 | 87 | 323 | 73.1 |
| HNSC | 20531 | 415 | 151 | 566 | 73.3 |
| LUSC | 20531 | 408 | 144 | 552 | 73.9 |
| MESO | 20531 | 71 | 16 | 87 | 81.6 |
| ESCA | 19076 | 165 | 31 | 196 | 84.2 |
| Total (PANCAN) |  | 4668 | 3213 | 7881 | 59.2 |

| **Pediatric Types** | **genes** | **Male** | **Female** | **Total** | **% Male** |
| --- | --- | --- | --- | --- | --- |
| Neuroblastoma | 22586 | 104 | 105 | 209 | 49.8 |
| Ependymoma | 25076 | 44 | 32 | 76 | 57.9 |
| High grade glioma | 25076 | 43 | 56 | 99 | 43.4 |
| Medulloblastoma | 25076 | 68 | 37 | 105 | 64.8 |
| Total Pediatric |  | 259 | 230 | 489 | 54.0 |

Next, we sought to define the transcriptomic sex index (TSI) as the Bayesian posterior probability of predicting a patient’s sex based on transcriptomic similarity to other samples. In our previous study we used the Joint and Individual Variance Explained (JIVE) approach to categorically assign transcriptomes to sex-specific subtypes of glioblastoma [22]. An advantage of the Bayesian Nearest Neighbor (BNN) algorithm is that it can infer “breakpoints” between local groupings of nearest neighbors and estimate individual TSI values for any transcriptome along a continuous spectrum of values as a Bayesian posterior probability using that transcriptomes’ local neighbors [24].

Unlike the popular K-Nearest Neighbors (k-NN) classification method, BNN does not require pre-defining the number of neighbors. Instead, the algorithm computes a posterior probability distribution for k given a target sample within a dataset, assuming that samples closer to the target are more likely to come from the same source, such as a cluster or subtype. The algorithm then calculates a final posterior probability for predicting the target sample by integrating over the distribution of the number of neighbors (k).

The TSI value, calculated using BNN posterior probability, helps us evaluate the enrichment of male or female samples among nearby neighbors with similar expression profiles. A value close to 1 indicates the vicinity is highly enriched with male samples, while a value close to 0 indicates enrichment with females.

### Downstream analysis

After estimating TSI for each patient, the association between TSI and gene expression was assessed. Genes with expressions positively associated with male TSI were identified as male-biased genes, while genes with expressions negatively associated with male TSI were identified as female-biased genes. Enrichment of male-/female-biased genes were tested in MSigDB hallmark gene sets.

## RESULTS

We previously applied the Joint and Individual Variance Explained (JIVE) algorithm to decompose male and female glioblastoma transcriptome data into components shared among males and females, and those unique to each sex [22]. This allowed us to identify “sex-specific” gene expression patterns and sex-based molecular subtypes of GBM. While informative, the JIVE approach may be limited in clinical utility because of the requirement for categorical assignment of gene expression to “male-specific”, “female-specific”, and shared components. “Sex” as a categorical variable has limited utility when investigating sex differences or attempting to stratify individual patients for sex-adapted treatments. This is because sex differences in cancer occur along a spectrum of phenotypes, like height, a critical feature that is not captured in the JIVE approach. Thus, we sought to develop a method for generating individual patient-specific values along an axis that traversed between male and female cancer transcriptional “poles”.

We created UMAPs from the transcriptomes of each cancer based on similarities in gene expression. The head and neck squamous carcinoma (HNSC) UMAP, illustrates the process of identifying male and female “poles” in the data (**Fig 1A**). We discovered local areas in the UMAP where samples were primarily male (blue circle) or female (red circle). We quantified the local sex enrichment and used them as poles to detect sex-bias in cancer transcriptomes. We defined TSI as the Bayesian posterior probability of predicting the sex status of a patient based on their individual transcriptomic similarity to the male or female poles. Thus, this index value reflects local transcriptomic patterns. A Bayesian algorithm [24] was used to infer “breakpoints” between local and remote samples. High TSI values are locally enriched in male cases, while low TSI values are locally enriched in female cases.

**Figure 1:**
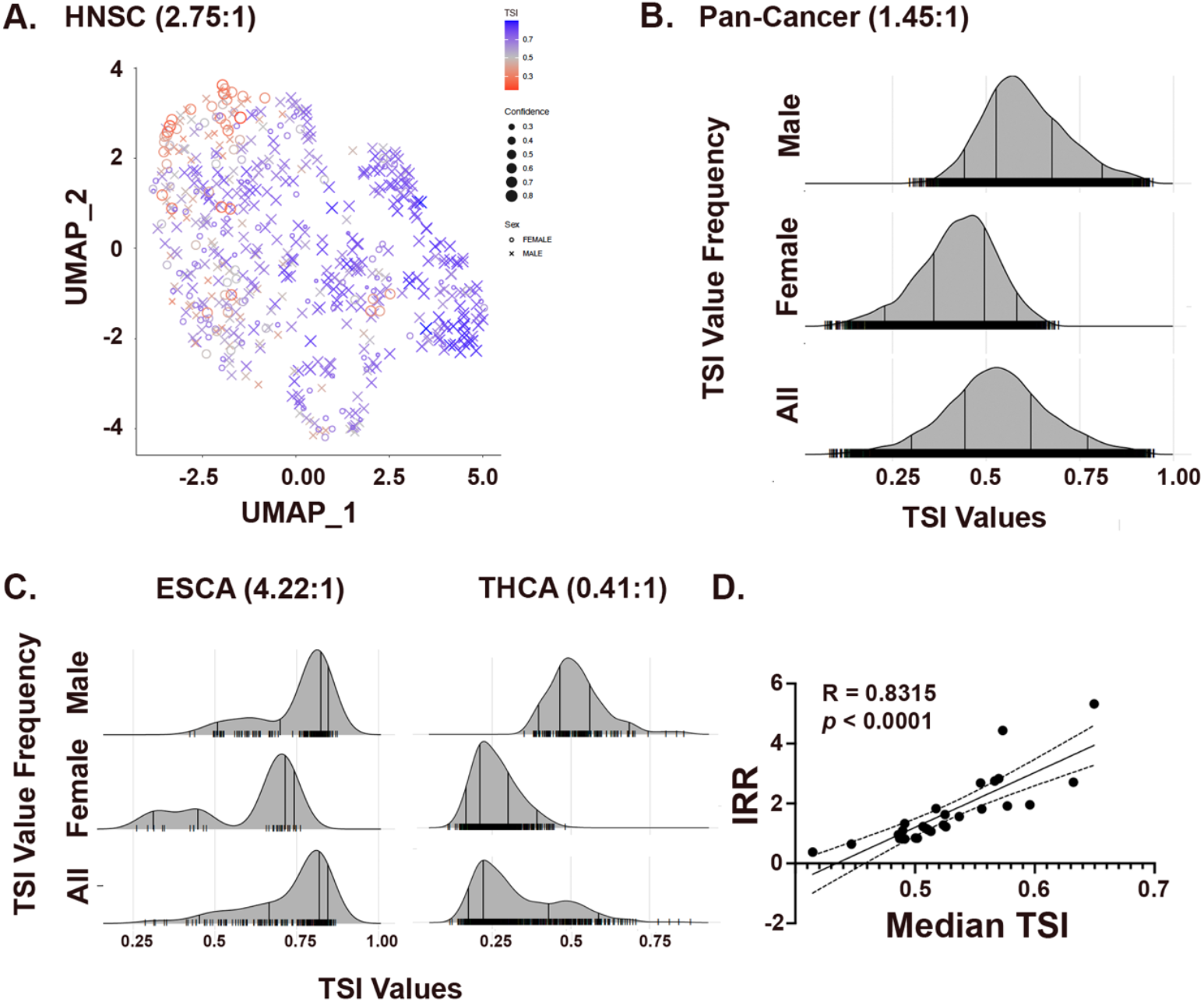
Cancer transcriptomes are sex-biased. (**A**) UMAP of 566 HNSC transcriptomes clustered by similarity. Male:Female Incidence rate ratio is shown. Male (blue X’s) and female (Red circles) distribute throughout the transcriptional space. Local enrichments for male and female transcriptomes were recognized and quantified to define male and female poles of gene expression. (**B**) Ridge plots of the TSI value distributions for Male, Female, and All patients from the PANCAN data (7881 total, 4668 M, 3213 F., M:F IRR = 1.45:1) (**C**) Ridge plots for TSI population distributions for Esophageal Carcinoma (ESCA, M/F IRR = 4.22) and Thyroid Carcinoma (THCA, M/F IRR = 0.7) illustrate the correlation between IRRs and median TSI values for the 26 adult cancers. (**D**) Regression analysis of IRR vs. Median TSI values. Shown is the best fit and 95% confidence intervals. R and *p* values are shown.

We derived TSI values for all individual cases and median TSI values in every cancer population separately (**Supplemental Figure 1)**. Then, we combined all 7881 adult TSI values in order to create a pan-cancer TSI population distribution (**Fig 1B**). As can be seen, the male and female values are skewed. Further, cases with TSI values below 0.25 are exclusively female and only males have values above 0.75. These cases represent the female and male population poles, respectively. It is also clear from the data that a large fraction of the cancer population possesses TSI values between 0.25 and 0.75. As female cases approach TSI values of 0.5, they represent changing balances between loss of female pole effects, and gain of male pole effects; while for males, there is loss of male-biased and gain of female-biased effects.

As expected, median TSI values for each cancer type positively correlated with their incidence rate ratios (IRR) as calculated from these datasets. As illustrated in **Fig 1C**, esophageal carcinoma (M/F IRR = 5.32) and thyroid carcinoma (M/F IRR = 0.38) exhibit median TSI values of (0.65) and (0.41), respectively. Similarly, the other cancers with M/F IRRs of less than 1 (sarcoma, adrenocortical carcinoma, diffuse large B cell lymphoma, thyroid carcinoma) exhibit median TSI values of less than 50 (**Supplemental Table 1**). Regression analysis of IRR versus median TSI identified a significant correlation between the two (**Fig 1D**). Thus, TSI value distributions are concordant with sex differences in individual cancer IRRs. Importantly, TSI values indicate that many males with female predominant cancers exhibit individual TSI values that are shifted towards the female pole and that many females with male predominant cancers exhibit individual TSI values that are shifted towards the male pole.

Together, these data suggest that sex effects on gene expression are pervasive and interact with cancer type-specific effects on gene expression to produce individual transcriptomes. Thus, we next sought to identify the genes and pathways that define the high and low TSI poles. We did so by looking for consistency in sex-biased mechanisms across cancer types using the 7881 adult and in parallel, the 1069 pediatric cases. Those genes with the greatest effect on low and high TSI values were identified by performing association analysis between TSI and gene expression. Genes that were significantly (FDR<0.05) associated with high male TSI were identified as “male-biased”, while those negatively associated with high TSI were identified as “female-biased” (**Supplemental Table 2**).

Cancer Hallmark Pathway analysis across cancer types indicated that most hallmark pathways [25] exhibit sex differences in gene expression and revealed several patterns of transcriptome polarization (**Figure 2A and B)**. Seventeen of the 26 cancer types were enriched for genes involved in oxidative phosphorylation and/or cell cycle regulation at the male pole. Twelve of the 26 cancers were enriched for genes involved in inflammation and immunity at the female pole (**Figure 2A and B**).

**Figure 2:**
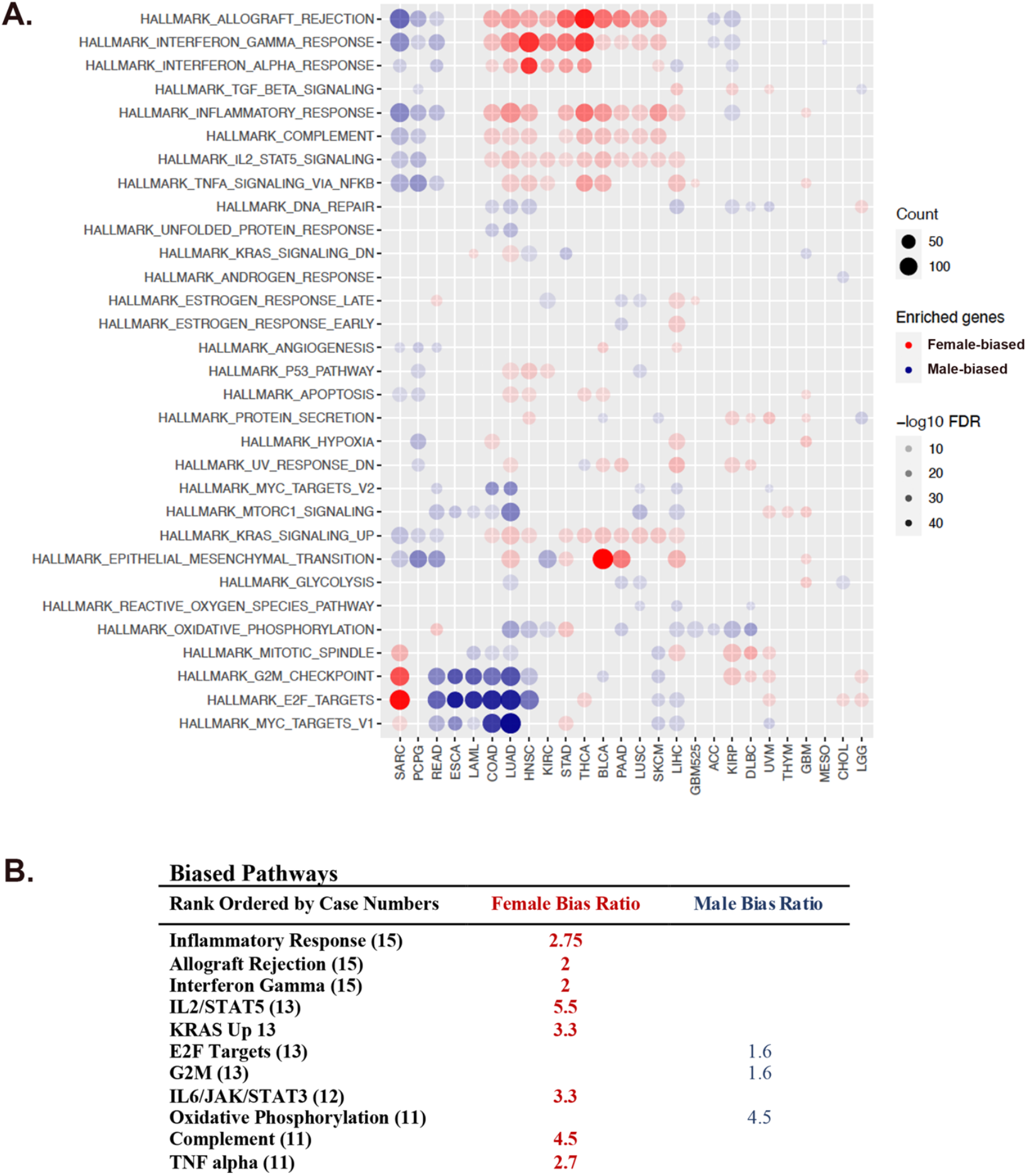
Most Cancers exhibit sex-biased cancer hallmark pathway activation. (**A**) Genes with the greatest effect on low (female pole) and high (male pole) TSI values were identified. Cancer Hallmark Pathway analysis of pole-associated genes revealed a predominant polarization pattern involving cell cycle regulation and oxidative phosphorylation at the male pole (blue circles) and multiple inflammatory/immunity pathways at the female pole (red circles). Gene counts (count) are symbolized by the size of the circles and False Discovery Rates (FDR) by the saturation of the fill as indicated in the legends. (**B**) The most commonly biased pathways listed in rank order of cases involved (in parentheses) and the ratio of female biased cases (red text) and male biased cases (blue text).

Thymoma was the only cancer without evidence of polarization. Interestingly, sarcoma differed from the predominant polarization patterns such that male cases were enriched for inflammation/immunity signatures and female cases for cell cycle regulation.

These data indicate that varying degrees of sex biased gene expression exist across cancer types and that a predominant shared pattern between multiple cancer involves biased gene expression in cell cycle regulation versus inflammation/immunity, pathways, which are known to be strongly sex-biased in action. Thus, we conclude that TSI values can successfully localize individual cancer cases along axes that traverse between sex-biased poles in targetable mechanisms like cell cycle regulation and immunity/inflammation.

As much of sexual differentiation is completed *in utero*, we hypothesized that similar sex differences in TSI would be evident in pediatric cancers. We applied the same strategy to the analysis of two pediatric transcriptome datasets: the Gabriella Miller Kids First Pediatric Research Program (Kids First (KF)), which included 209 neuroblastoma patients and the Children’s Brain Tumor Network (CBTN), which included 865 patients comprised of 101 high grade glioma, 105 medulloblastoma, 79 ependymoma, and 214 low grade glioma cases (**Table 1**). From the KF data, 6330 male biased-genes and 6089 female biased genes were identified (FDR<0.05, **Supplemental Table 3**). From the CBTTC data, 3063 male-biased genes and 2062 female-biased genes were identified (FDR<0.05, **Supplemental Table 4**). There were 1126 shared male-biased genes, and 742 shared female-biased genes between the two datasets (both with p<2.2e-16, **Supplemental Table 5**).

Like the adult cancers, these pediatric cancers exhibited biased distributions of TSI values (**Figure 3, Supplemental Figure 2)**. Neuroblastoma (M/F IRR = 1.11) is strongly polarized and again, those cases with high TSI values were enriched for cell cycle regulation while those associated with low TSI values were enriched for inflammation and immunity (**Figure 3B**). The CBTN brain tumor data includes the diverse histologies common in pediatric neuro-oncology. Ependymoma (M/F IRR = 1.5) exhibited the strongest polarization, in which low TSI values were enriched for inflammation and immunity and oxidative phosphorylation was strongly correlated with high TSI values (**Figure 3C**). The most common malignant brain tumor of childhood is medulloblastoma (M/F IRR: 1.8:1) [26]. The strongest association in medulloblastoma was between low TSI values and cell cycle regulation, reminiscent of what was observed for adult sarcomas. In pediatric high-grade glioma (M/F IRR ≈ 1), high TSI values were strongly correlated with cell cycle regulation, while there were no distinct gene expression patterns associated with low TSI value cases. Thus, like adult cancers, pediatric cancers exhibit sex-bias in gene expression that varies in magnitude and involved pathways, indicating that sex and tumor type interact to yield different TSI profiles in both adult and pediatric cancers. Importantly, sex biases in gene expression and pathway activation are evident even when the incidence ratios of particular cancer types, e.g., pediatric high-grade glioma, near equivalence. Therefore, individuals with any cancer type may be more extensively phenotyped for personalized approaches to treatment using a TSI analysis than without.

**Figure 3:**
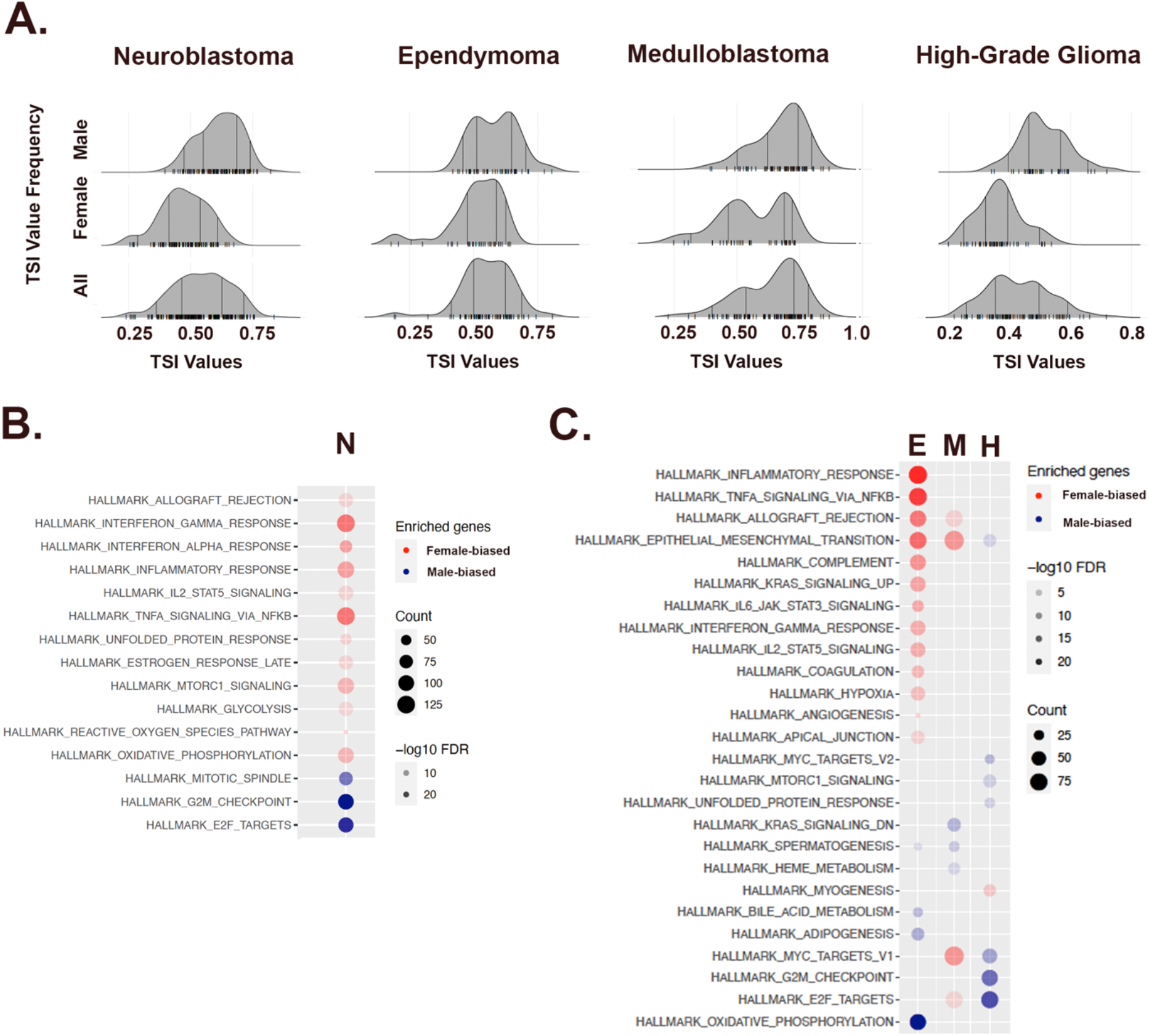
Pediatric neural tumors also exhibit sex biases in gene expression. (**A**) Ridge plots for neuroblastoma and the three most common malignant brain tumors of childhood (489 total, 259 M, 230 F., M:F IRR = 1.13:1)demonstrating sex biases in TSI population distributions. (**B**) Cancer Hallmark Pathway analysis of those genes that exerted the greatest effects on the male and female poles for each cancer. A predominant polarization pattern is identified with inflammatory/immunity pathways associated with the female pole (red circles) and cell cycle regulatory pathways associated with the male pole (blue circles). Gene counts (count) are symbolized by the size of the circles and False Discovery Rates (FDR) by the saturation of the fill as indicated in the legends.

Cancer patients with extremes of high and low TSI values might be approachable with something akin to sex-specific treatments as they do not exhibit measurable influence of the opposite sex-bias in gene expression. However, TSI values for most patients lie between the poles. Therefore, we expected that their transcriptomes would exhibit both male- and female-biased components. We hypothesized that for female cases, translation along the TSI axis from the female pole (TSI < 0.25) to midrange values would involve decreased female-biased effects and/or increased male-biased effects. We predicted that the opposite would be the case for male cases with midrange TSI values compared to the male pole (TSI > 0.75). If this proved to be the case, we expected this could serve as a tool for stratification for sex-adapted treatment planning, even for male and female patients with identical TSI values. To address this hypothesis, we first compared the PANCAN transcriptomes of male or female cases with midrange TSI values to those with transcriptomes closer to their respective poles. We then performed pathway analysis to determine which pathways were altered relative to the poles. Most female cases exhibited a loss of the inflammatory/immunity signatures (**Figure 4A**). Four cases (LUAD, LIHC, COAD, DLBC) exhibited a gain in a male cell cycle regulatory signature as well as a male epithelial-to-mesenchymal signature. More male mid-range cases exhibited concomitant transcriptional changes. For many male cases, there was a clear increase in the inflammation/immunity signature characteristic of the female pole (**Figure 4A**). There was also a mix of cases with decreased (COAD, LUAD, LIHC, LAML, READ, ESCA, PCPG) or increased (HNSC, STAD, LUSC, LUSC, BLCA, KIRP, UVM), male cell cycle regulatory signatures. Interestingly, male sarcoma cases with midrange TSI values did not exhibit changes in pathway activation signatures that differed from the male pole, suggesting the possible involvement of other factors that favor lower TSI values. Together these data emphasize the potential of this approach for identifying sex-biased actions underlying mid-range TSI values, as the TSI approach highlights sex biased-mechanisms for those patients at any point on the TSI scale.

**Figure 4:**
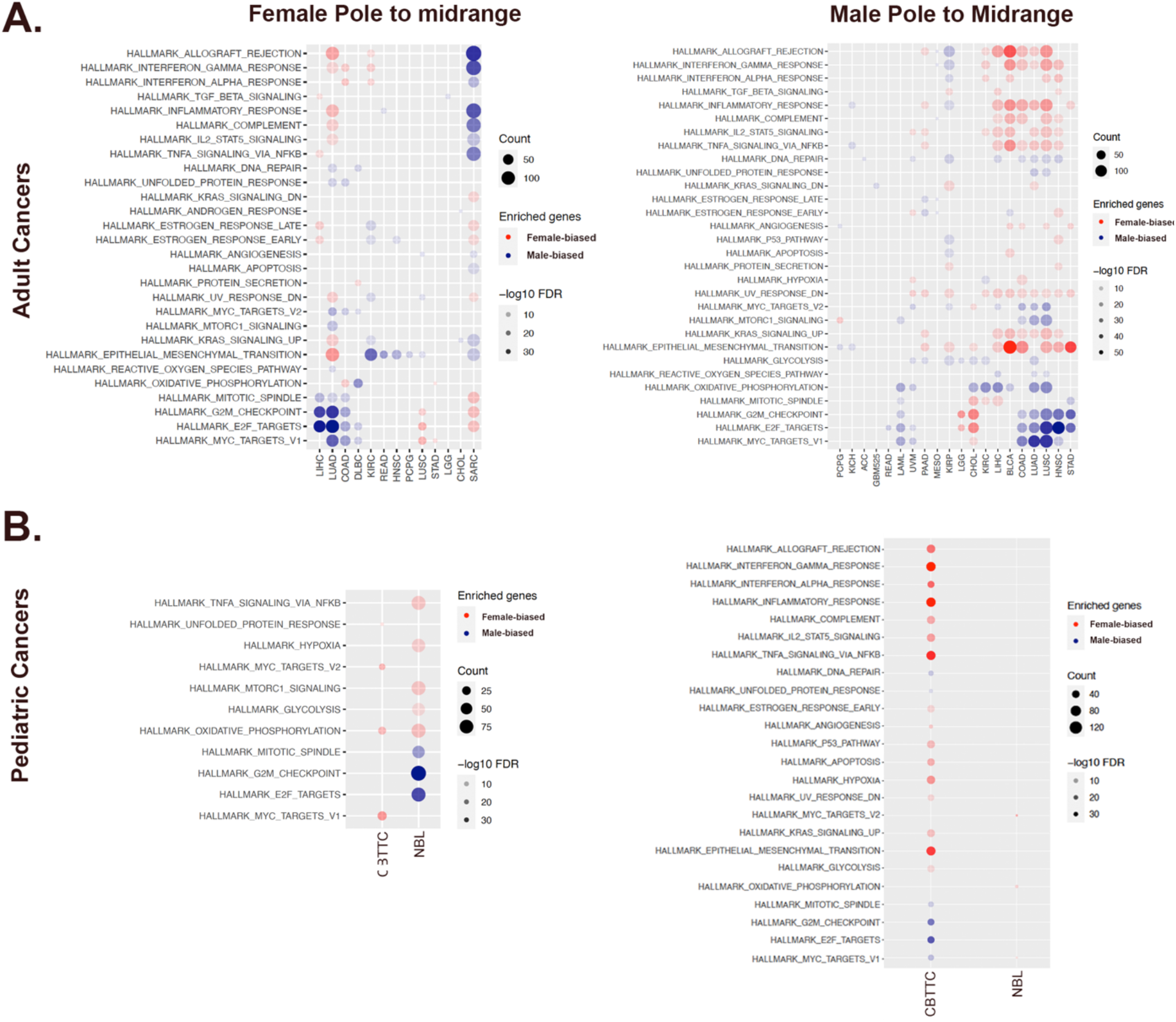
Mid-range TSI values exhibit distinct pathway signatures relative to the sex-specific poles. (**A**) (Left Panel) Heatmap of pathway activation signatures underlying changes in midrange TSI values for female adult cancers relative to their pole. (Right Panel) Heatmap of pathway activation signatures underlying changes in midrange TSI values for male adult cancers relative their pole. (**B**) (Left Panel) Heatmap of pathway activation signatures underlying changes in midrange TSI values for pediatric female cancers relative to their pole. (Right Panel) Heatmap of pathway activation signatures underlying changes in midrange TSI values for pediatric male cancers relative their pole. For all panels, changes in male (blue circles) and female (red circles) signatures are indicated. Gene counts (count) are symbolized by the size of the circles and False Discovery Rates (FDR) by the saturation of the fill as indicated in the legends.

Next, we performed the same analysis in the pediatric datasets. Similar to what we found for the PANCAN data, translation away from their respective poles to midrange TSI values occurred concomitantly with a shift in pole-defining pathway involvement (**Figure 4B**). In the CBTTC data, we found that females with midrange TSI values exhibited decreased MYC targets (V1 and V2) and oxidative phosphorylation without gain of male biased pathway activation. For males with midrange TSI values in the CBTTC data, we saw significant changes in almost all hallmark pathways with gains in both the female inflammatory signature as well as gains in a male cell cycle regulatory signature, much as we had seen in the PANCAN male cases with midrange TSI values. In addition, a number of other pathways demonstrated a shift towards a female bias. In the KF Neuroblastoma data, females with midrange TSI values exhibited a strong acquisition of a male cell cycle regulatory signature as well as a decreased female inflammation/immunity and metabolism signatures (**Figure 4B**). No biased pathway signature changes were detectable in the male midrange TSI value cases. Together, these results support the hypothesis that midrange TSI values are associated with different molecular pathway activation profiles for males and females with different cancer diagnoses. Thus, even when similar in phenotype, particular subsets of male and female cancer patients may benefit from sex-adapted therapies.

## Discussion

Sex effects in cancer incidence, treatment, and survival are commonplace [2]. Like any other significant difference in cancer phenotypes, understanding the mechanistic basis for sex differences holds promise for improving outcomes for all. Sex-adaptations to treatment are complicated by the nature of sex differences. Most sex differences are not as dichotomous as a peacock’s tail. Instead, individuals are unique mixes of maternal and paternal features, and a singular balance between male- and female-biased actions. This suggests that the optimal translation of sex differences in cancer biology will require recognizing sex effects as continuous variables and making measurements of the individual mix of male- and female-biased actions in each patient.

Here, we showed that relative male- and female-biased actions can be quantified through a Bayesian Nearest Neighbor-based derivation of a Transcriptomic Sex Index. The value of this approach is it uses the entire patient-specific tumor transcriptome, as well as a realistic treatment of sex differences as continuous variables, to make predictions regarding the relative contributions that targetable male- and female-biased actions make to each individual patient’s cancer biology.

To do this, we first identified male and female extremes in cancer transcriptional phenotypes and then, the genes and pathways underlying these poles. We observed several different polarization patterns, highlighting that sex interacts in variable ways with differing cancer mechanisms, their cells of origin and oncogenic events, their tissue and systems biology. The most common polarizations occur around male - cell cycle regulation and female - inflammation/immunity in both adult and pediatric cancers. This serves as validation for this approach as both mechanisms are already known to exhibit male- and female-biased action under normal conditions [11-14]. As both pathways are targetable with available therapeutics [27, 28], it is interesting to consider how the TSI approach might inform stratification for treatment. As an illustration, clinical experience indicates that females exhibit a smaller survival benefit from immune checkpoint inhibition (ICI) than males [29-31]. Thus, it would be reasonable to hypothesize that the immune signature associated with the lowest TSI values is one of resistance to ICI. If so, then females with low TSI values would be less responsive to this treatment compared to females with higher TSI values. Concordantly, males have been shown to be more responsive to ICI and therefore, high TSI values are a biomarker for sensitivity to ICI, while males with lower TSI values may be less so (**Figure 5**). In this way, sex, cancer type, and TSI values might more precisely stratify patients for ICI, or by analogy other targetable pathways with recognizable sex differences.

**Figure 5:**
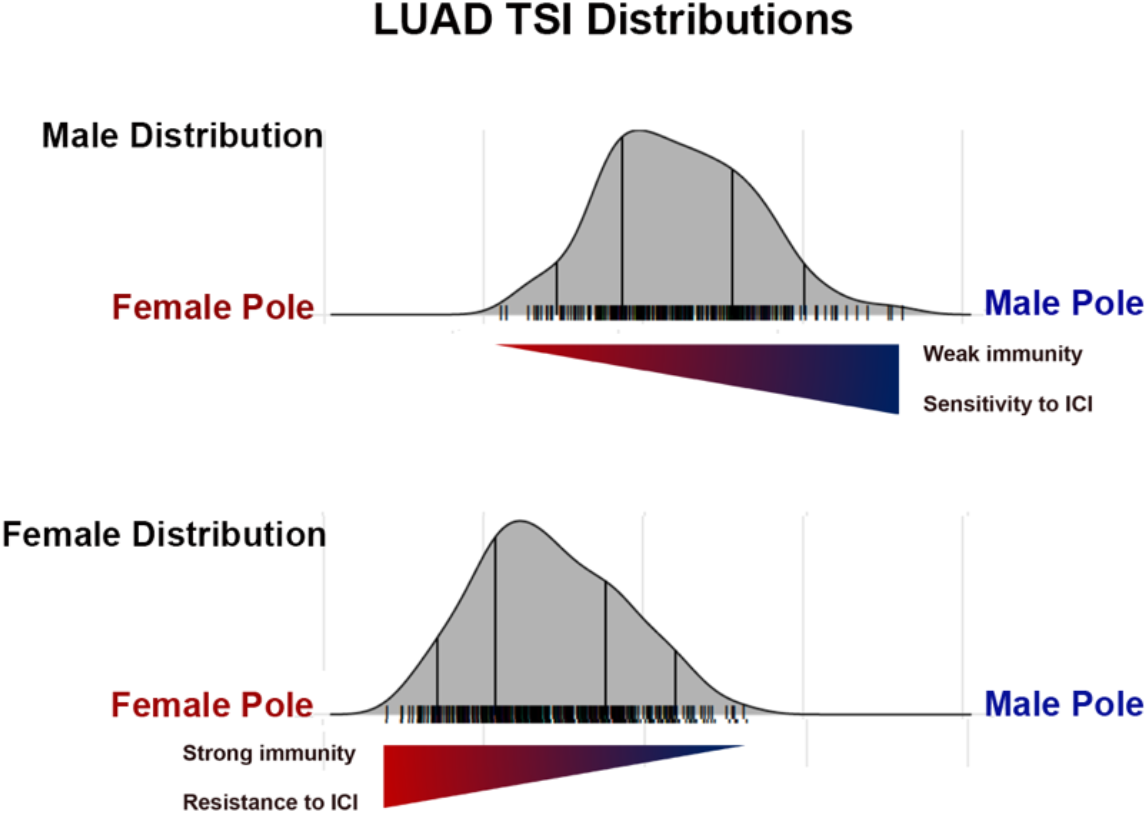
Example application of TSI in patient treatment stratification. Pictured are the TSI distributions for female (top panel) and male (bottom panel) lung adenocarcinoma (LUAD) patients. Females exhibited a strong immunity/inflammation signature and in clinical trials, are resistant to immune checkpoint inhibition (ICI). In contrast, male LUAD patients do not exhibit an inflammation/immunity signature and are responsive to ICI. If male and female patients were stratified for immune checkpoint inhibition treatment, the most likely males to respond to treatment would be those with the highest TSI, those nearest the male pole. Female patients most likely to respond to treatment would also be those with the highest TSI values, those furthest from the female pole.

In conclusion, we can no longer focus on questioning whether or not biological sex differences matter in cancer. Sex differences in incidence, response to standard treatments, and survival, all strongly argue that they do. The question now is, how do we use the continuously varying nature of sex differences for treatment planning, patient stratification, and analysis of laboratory and clinical research results. As the TSI analysis enriches patient-specific phenotyping, we expect that its use can enhance personalized approaches to treatment by realistically accounting for sex effects in cancer.

Finally, The TSI approach would support moving away from terms like “sex-biased” and towards the use of low and high TSI values as they pertain to specific cancer types and the range of TSI values that occur in specific targetable pathways like cell cycle regulation, DNA repair, amino acid metabolism, inflammation, etc. This would reduce any ambiguity that arises from discussing male-biased mechanisms in female patients or female-biased mechanisms in male patients.

## Supporting information

Pole-defining genes for each cancer

Median TSI values for each cancer

Ridge plots for all pediatric cancers

Ridge plots for all adult cancers

shared_KF_CBTTC.gene_assoc..genes_group1.fdr0.05

cbttc_lm_strat.CBTTC.gene_assoc..genes_all.fdr0.05

KF.gene_assoc..genes_all.fdr0.05

## Data Availability

All data produced in the present work are contained in the manuscript

## Code availability

The R package for the Bayesian Nearest Neighbors method could be accessed from https://github.com/wuwill/BKNN

## Acknowledgements

Work on sex differences in the Rubin lab is supported by the National Cancer Institute R01 CA174737-06, P01CA245705, Joshua’s Great Things, Siteman Investment Program, Barnard Research Fund, St. Louis Children’s Hospital Foundation, Taylor Rozier’s Hope for a Cure Foundation, Prayers from Maria Foundation, and the Haubrich and Griffiths family foundations.

## Dataset acknowledgements

The results analyzed and published here are based upon datasets generated by the TCGA Research Network https://www.cancer.gov/tcga, accessed from https://pancanatlas.xenahubs.net, by Gabriella Miller Kids First Pediatric Research Program projects (phs001436.v1.p1), accessed from dbGaP (www.ncbi.nlm.nih.gov/gap), and by the Childhood Brain Tumor Tissue Consortium, accessed from https://cbtn.org.

## Notes

### Competing Interest Statement

The authors have declared no competing interest.

### Author Declarations

The results analyzed and published here are based upon datasets generated by the TCGA Research Network https://www.cancer.gov/tcga, accessed from https://pancanatlas.xenahubs.net, by Gabriella Miller Kids First Pediatric Research Program projects (phs001436.v1.p1), accessed from dbGaP (www.ncbi.nlm.nih.gov/gap), and by the Childhood Brain Tumor Tissue Consortium, accessed from https://cbtn.org. The use of publicly accessible human datasets for research has been approved by the Washington University Institutional Review Board (IRB# 201102299).

