## Supplementary material for "Accounting for sex differences variability in the design of sex-adapted cancer treatments": Median TSI values for each cancer

| **Supplemental Table 1** | | |
| --- | --- | --- |
| **Cancer Type** | **Median TSI value** | *IRR (M:F)* |
| ACC | 0.446836316 | 0.64583333 |
| BLCA | 0.55478254 | 2.68103448 |
| CHOL | 0.486033964 | 0.95652174 |
| COAD | 0.489726699 | 1.09361702 |
| DLBC | 0.501893389 | 0.84615385 |
| ESCA | 0.649604989 | 5.32258065 |
| GBM | 0.555716607 | 1.81355932 |
| HNSC | 0.566581024 | 2.74834437 |
| KICH | 0.491462713 | 1.33333333 |
| KIRC | 0.577054813 | 1.91346154 |
| KIRP | 0.632303448 | 2.71264368 |
| LAML | 0.509463259 | 1.1625 |
| LGG | 0.525722105 | 1.22268908 |
| LIHC | 0.595854361 | 1.95804196 |
| LUAD | 0.500412024 | 0.85209003 |
| LUSC | 0.569837159 | 2.83333333 |
| MESO | 0.573276386 | 4.4375 |
| PAAD | 0.506713549 | 1.23170732 |
| PCPG | 0.49149927 | 0.81553398 |
| READ | 0.510996242 | 1.125 |
| SARC | 0.487288126 | 0.82758621 |
| SKCM | 0.525150852 | 1.62777778 |
| STAD | 0.517501632 | 1.83018868 |
| THCA | 0.414638951 | 0.37831325 |
| THYM | 0.513283222 | 1.06779661 |
| UVM | 0.52376891 | 1.28571429 |
| GBM525 | 0.53696888 | 1.56097561 |
