## Supplementary material for "Accounting for sex differences variability in the design of sex-adapted cancer treatments": Ridge plots for all pediatric cancers

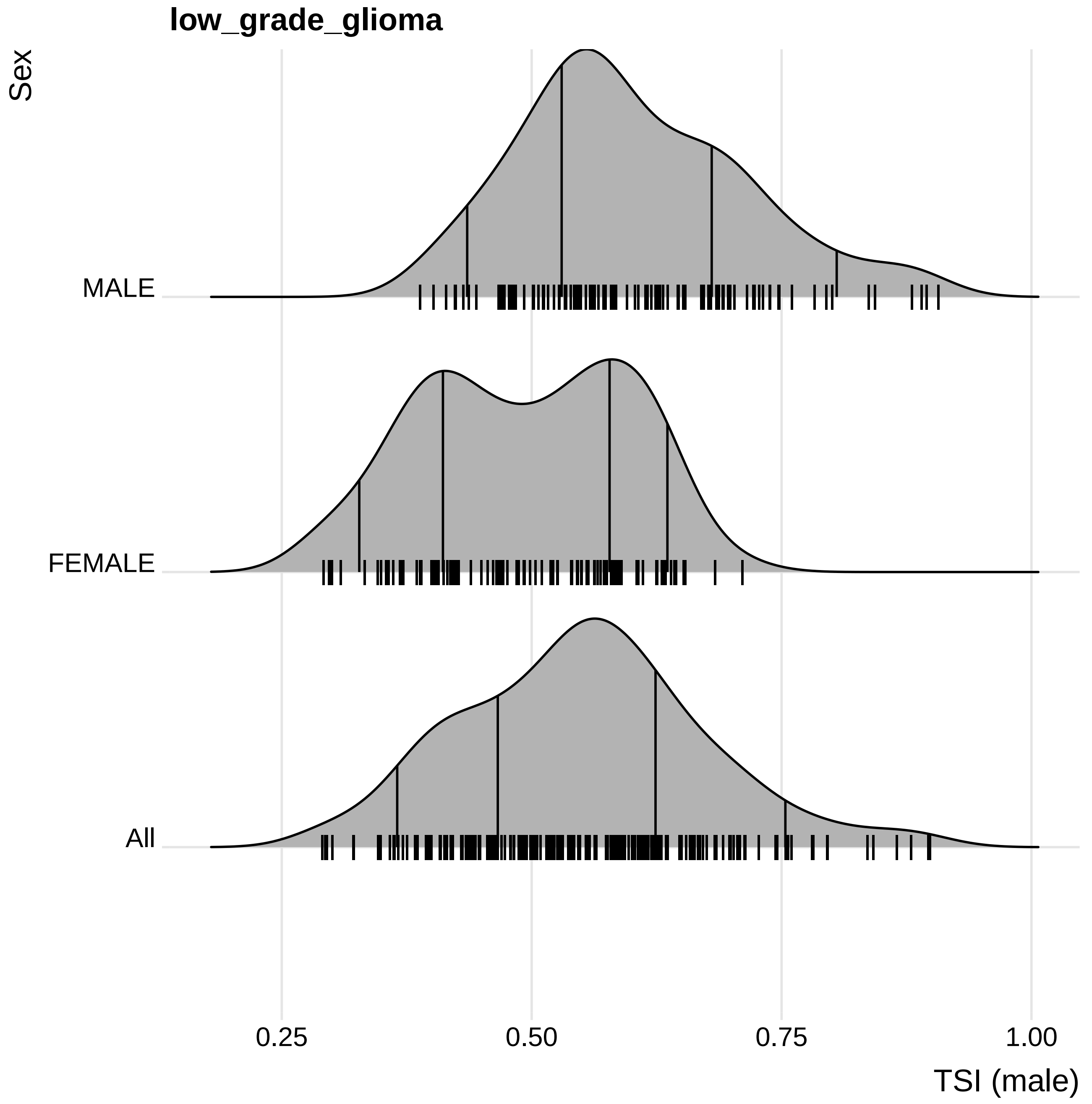
low_grade_glioma
Sex
MALE
FEMALE
All
0.25
0.50
0.75
1.00
TSI (male)

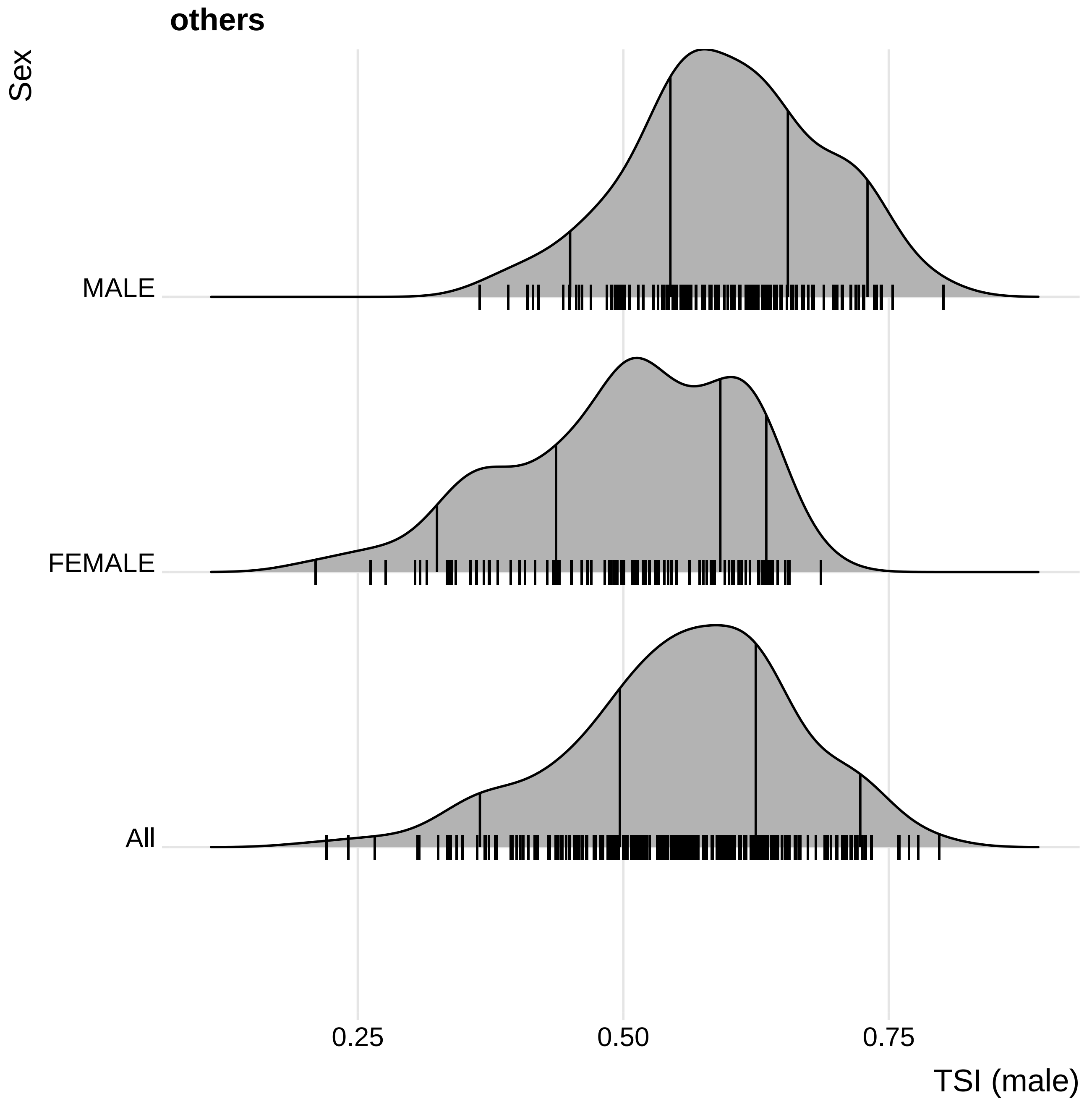

others
Sex
MALE
FEMALE
All
0.25
0.50
0.75
TSI (male)

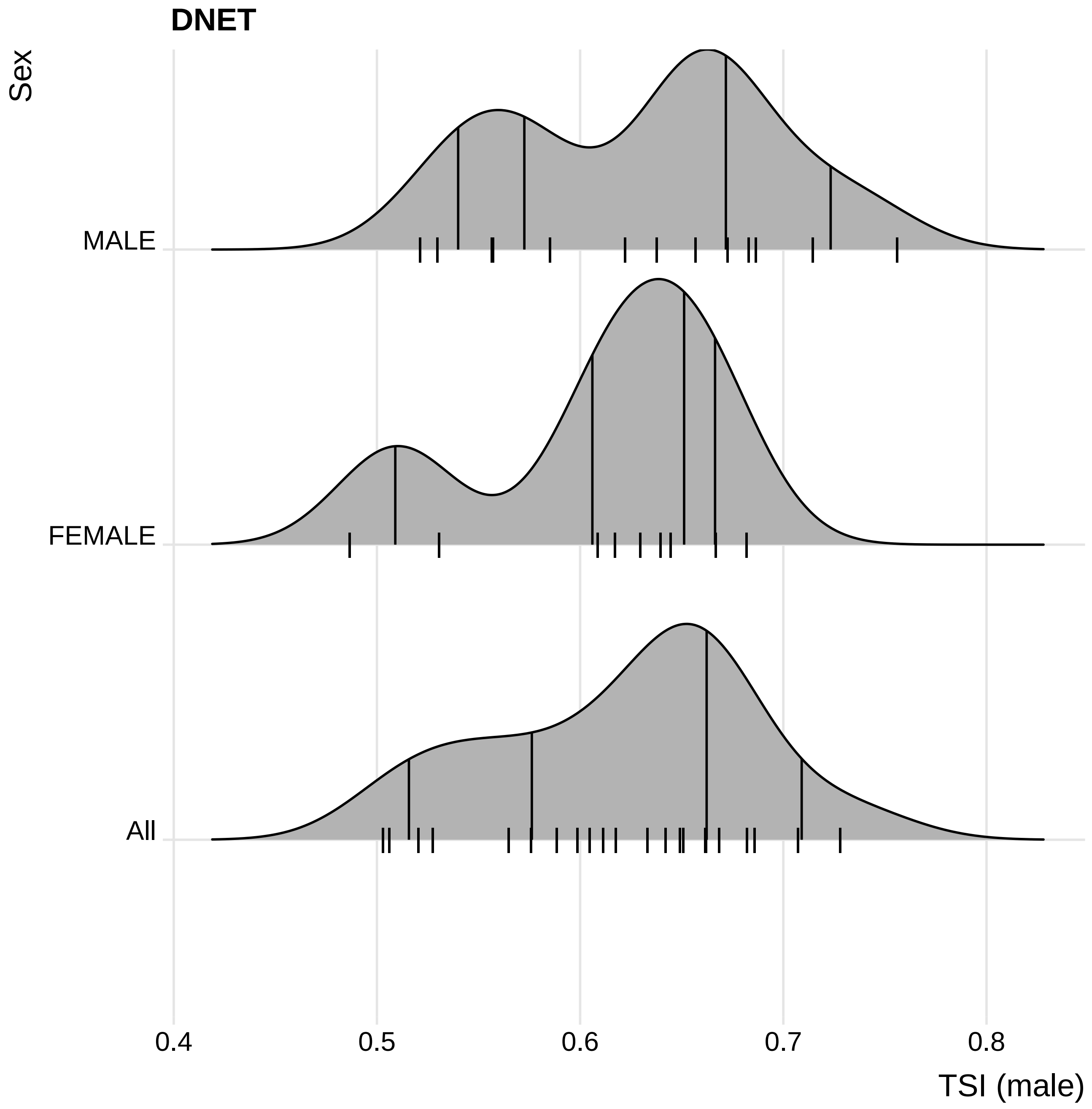
DNET
Sex
MALE
FEMALE
All
0.4
0.5
0.6
0.7
0.8
TSI (male)

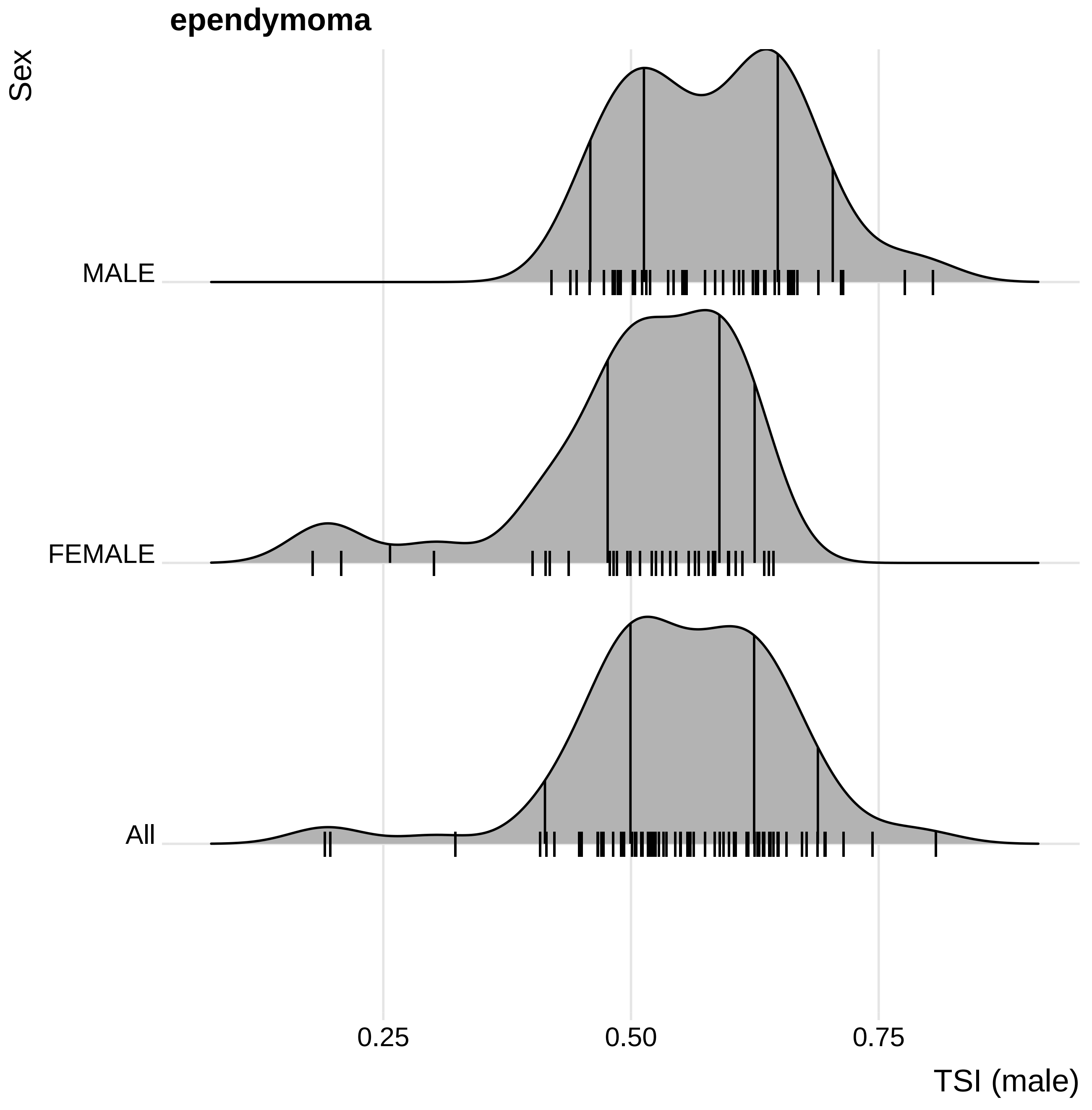

ependymoma
Sex
MALE
FEMALE
All
0.25
0.50
0.75
TSI (male)

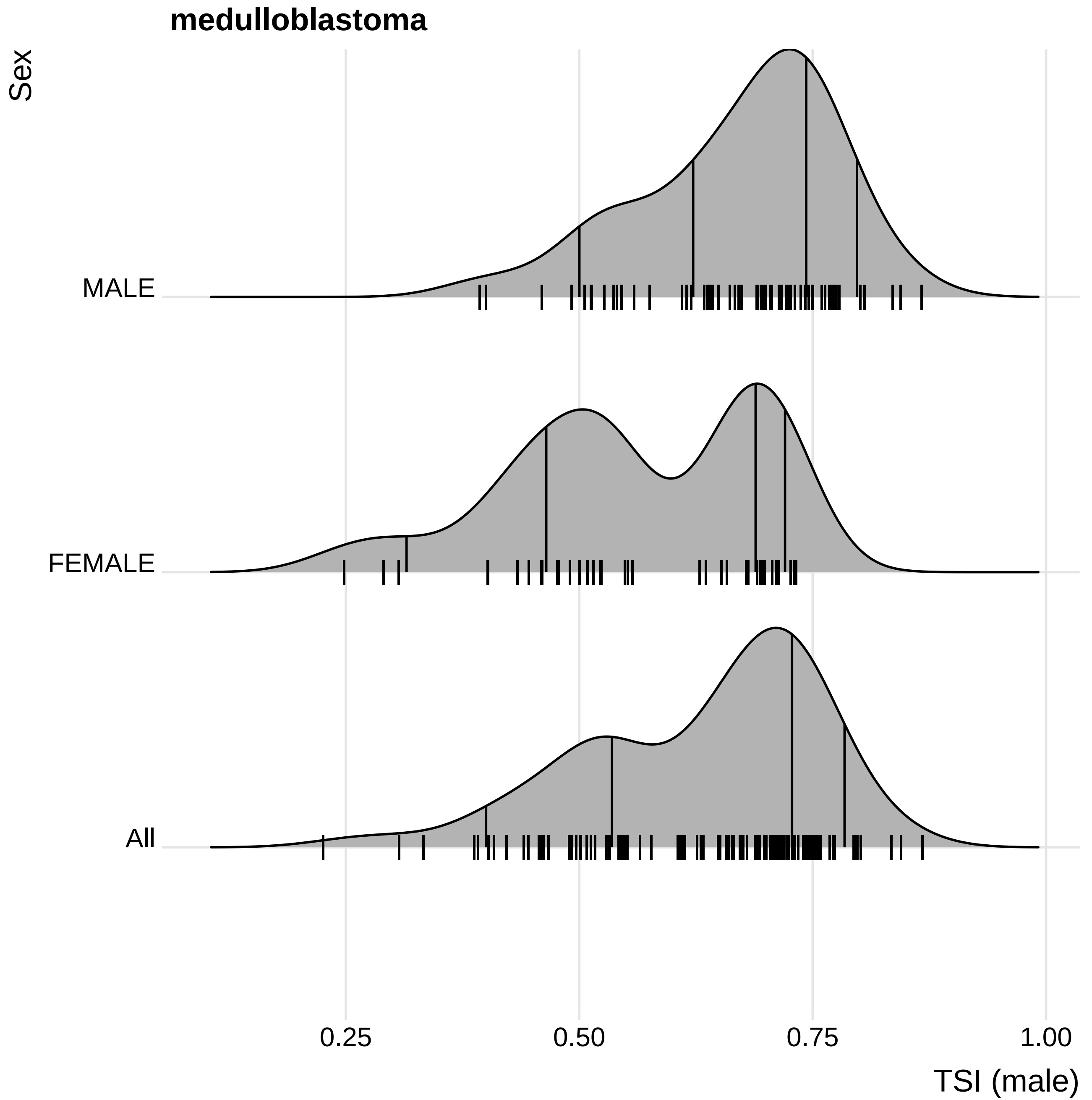
medulloblastoma
Sex
MALE
FEMALE
All
0.25
0.50
0.75
1.00
TSI (male)

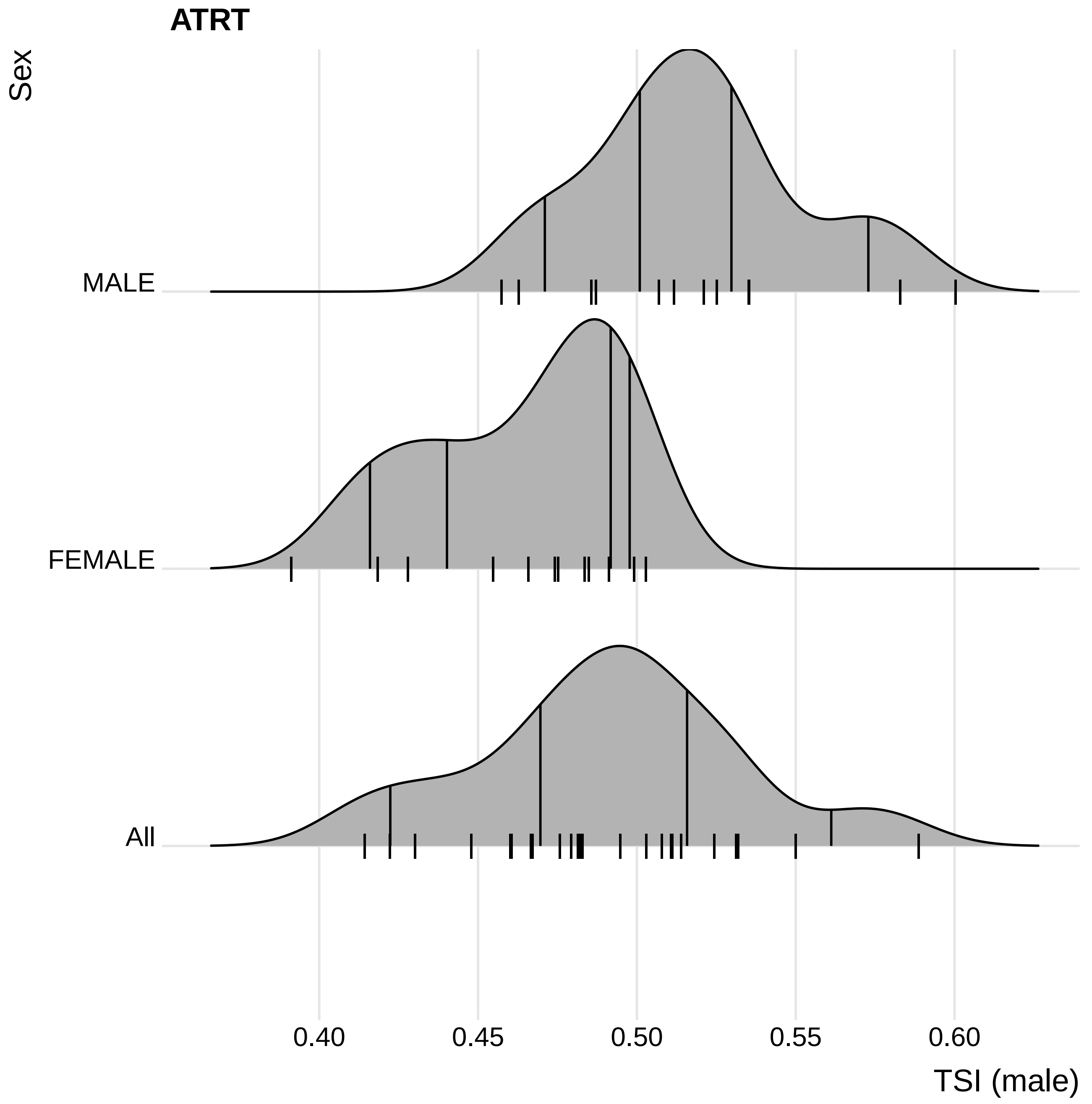
ATRT
Sex
MALE
FEMALE
All
0.40
0.45
0.50
0.55
0.60
TSI (male)

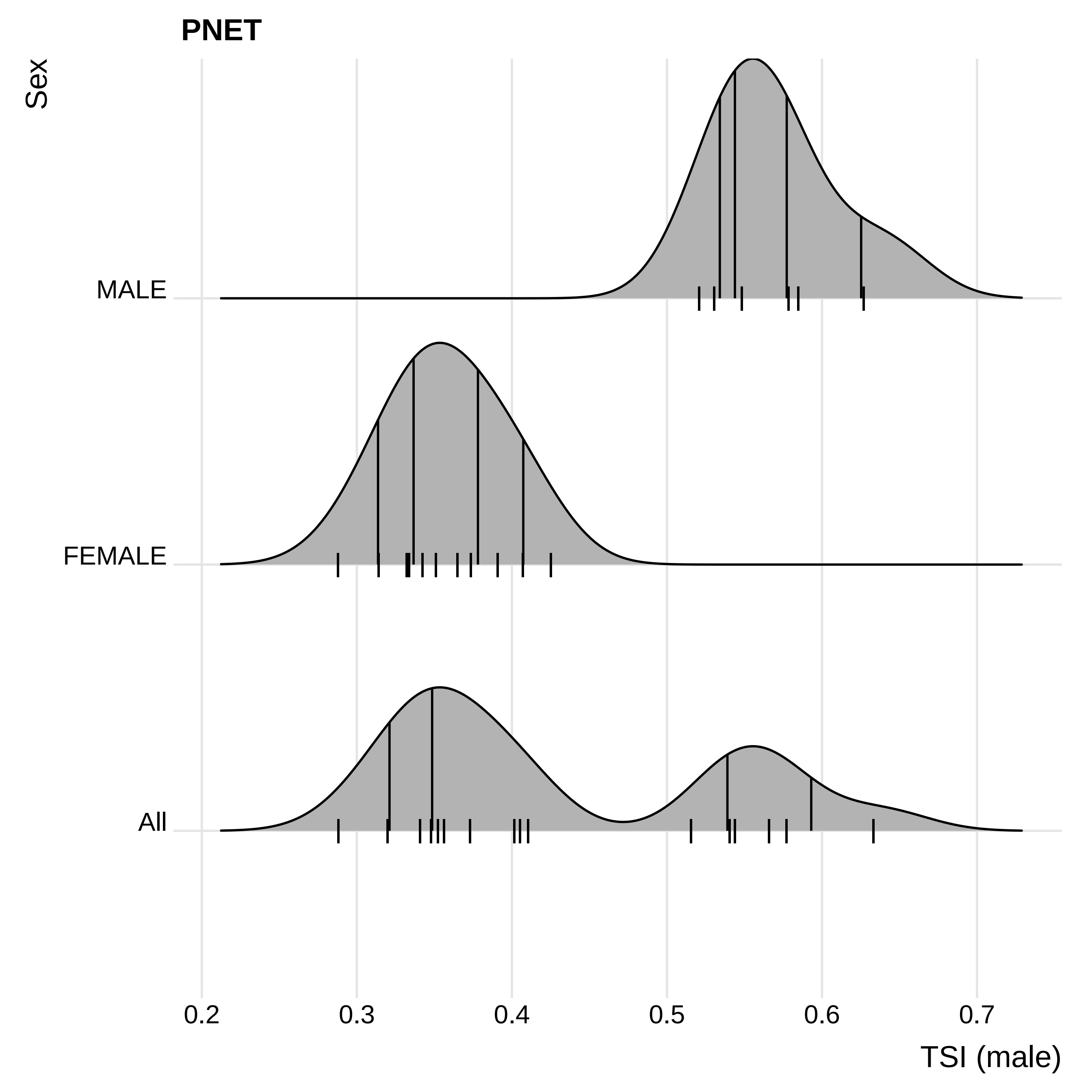
PNET
Sex
MALE
FEMALE
All
0.2
0.3
0.4
0.5
0.6
0.7
TSI (male)

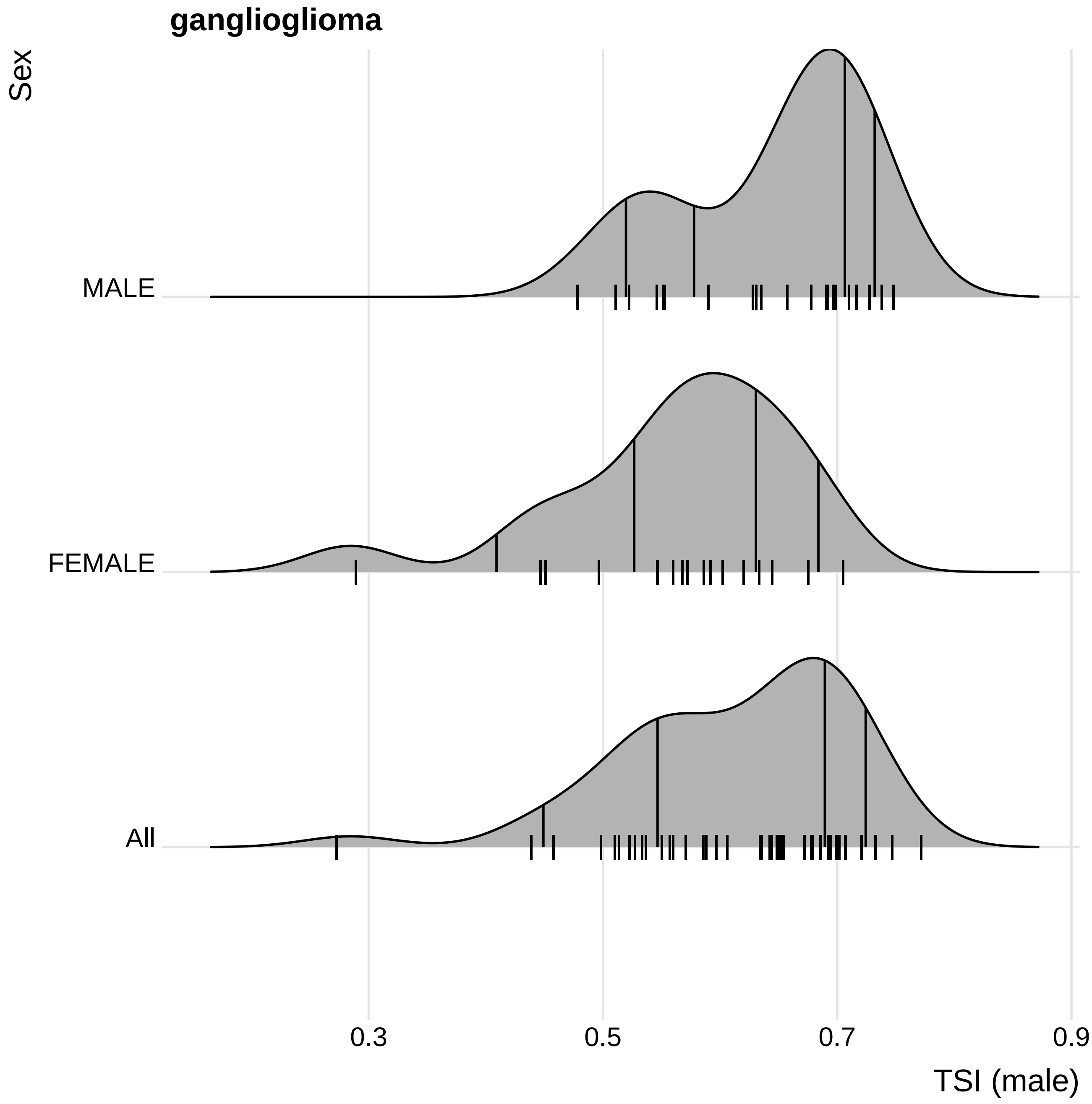
ganglioglioma
Sex
MALE
FEMALE
All
0.3
0.5
0.7
0.9
TSI (male)

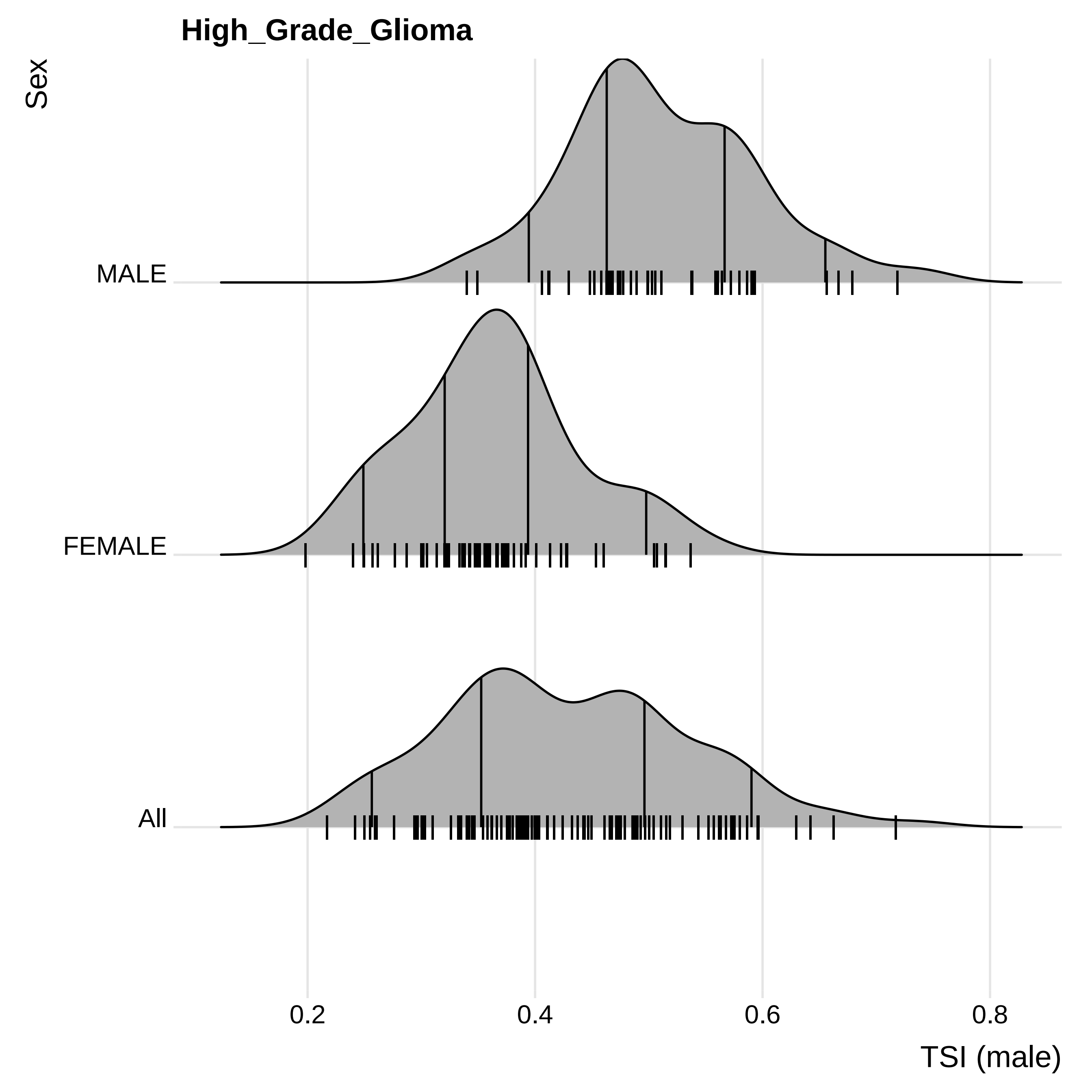

High_Grade_Glioma
Sex
MALE
FEMALE
All
0.2
0.4
0.6
0.8
TSI (male)

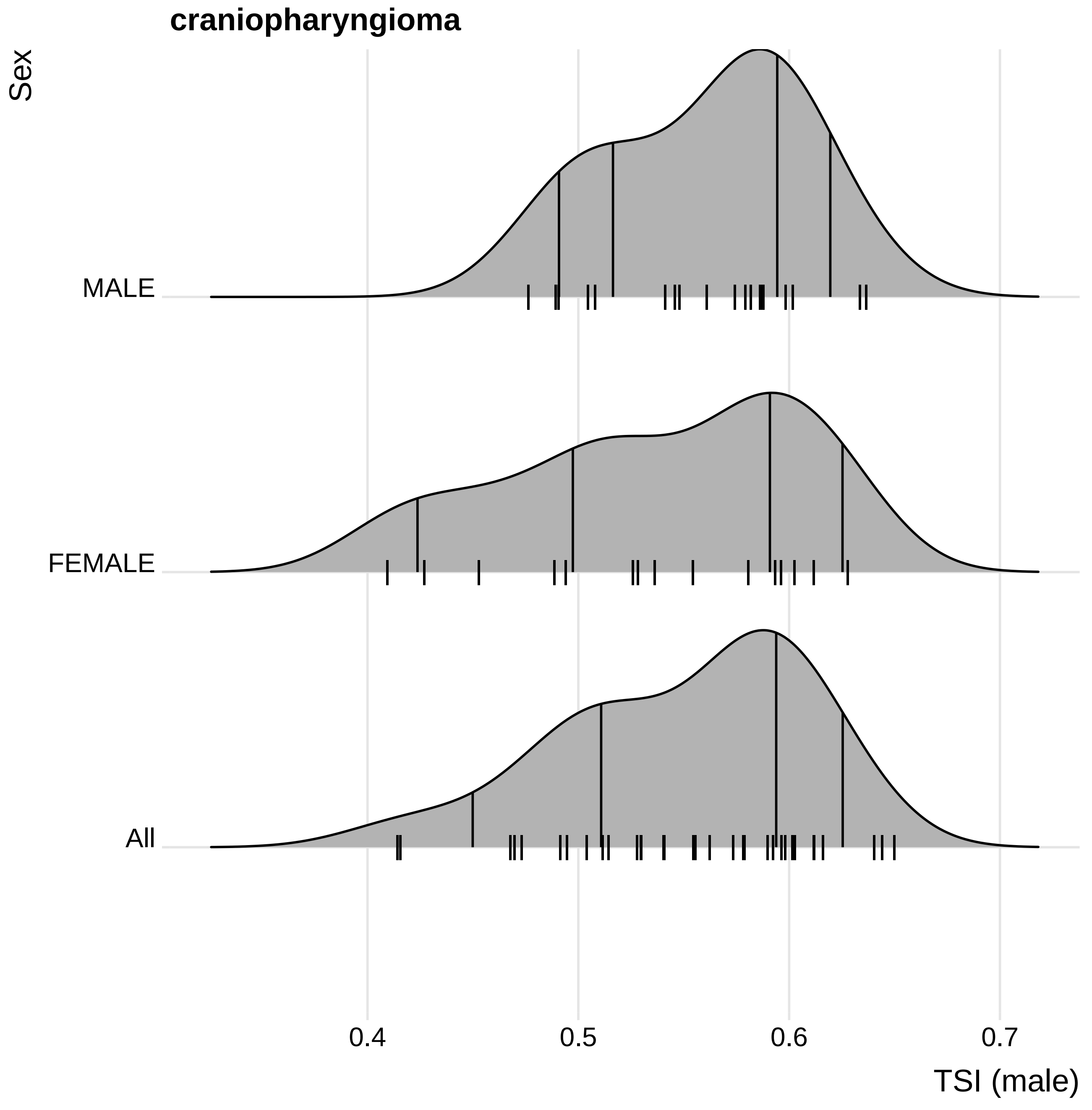
craniopharyngioma
Sex
MALE
FEMALE
All
0.4
0.5
0.6
0.7
TSI (male)

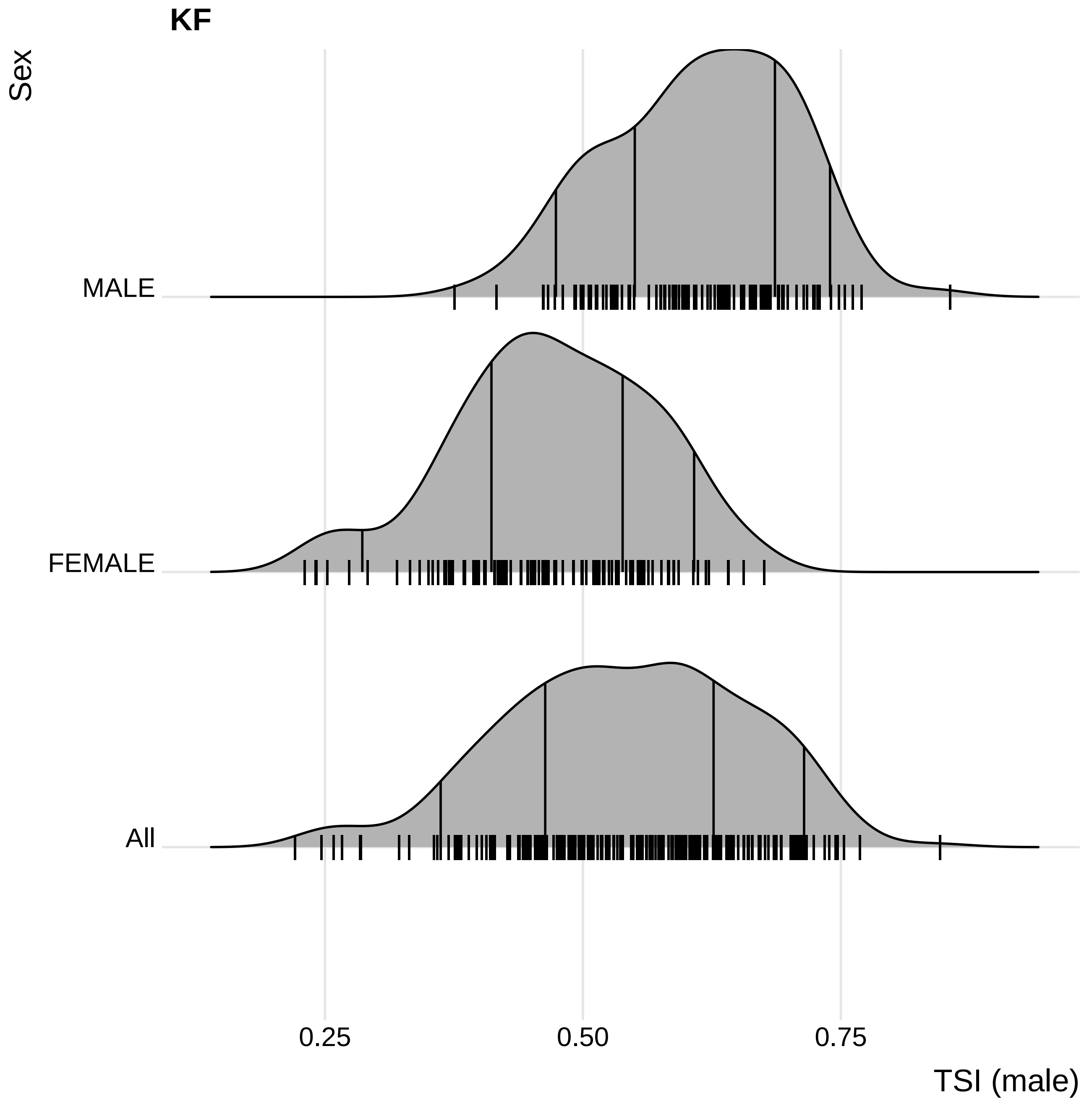

KF
Sex
MALE
FEMALE
All
0.25
0.50
0.75
TSI (male)
