## Supplementary material for "Accounting for sex differences variability in the design of sex-adapted cancer treatments": Ridge plots for all adult cancers

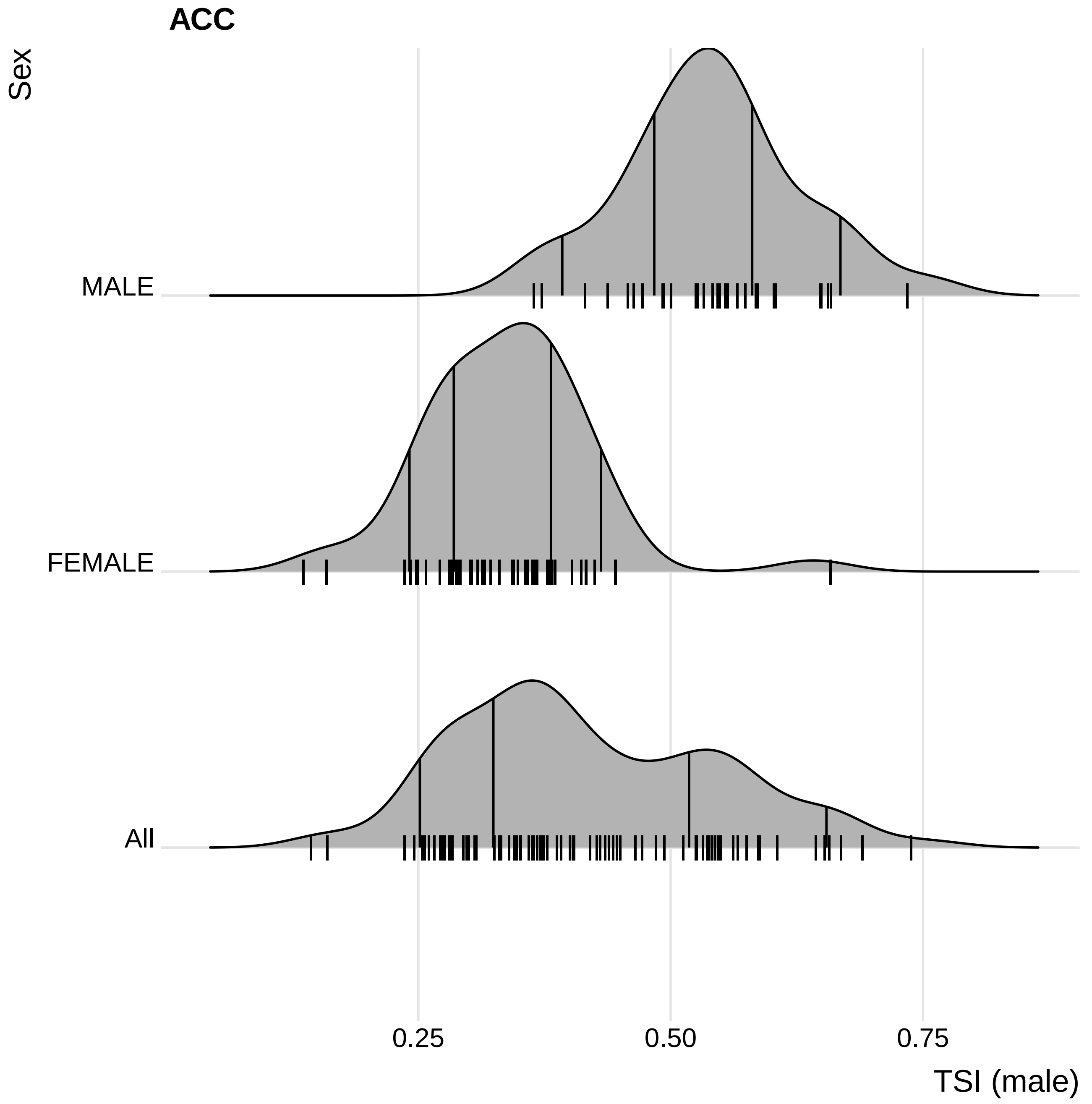
ACC
Sex
MALE
FEMALE
All
0.25
0.50
0.75
TSI (male)

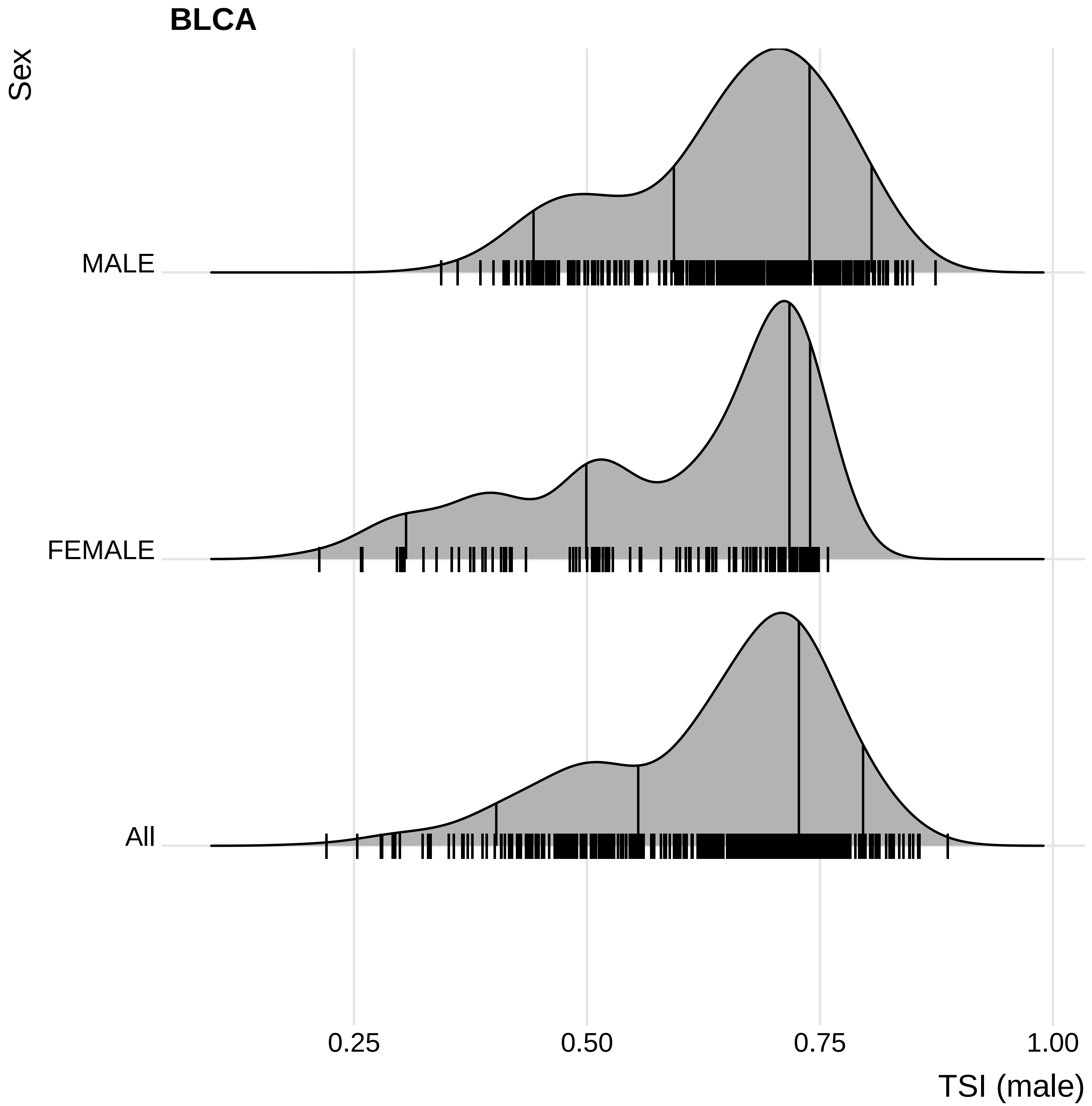

BLCA
Sex
MALE
FEMALE
All
0.25
0.50
0.75
1.00
TSI (male)

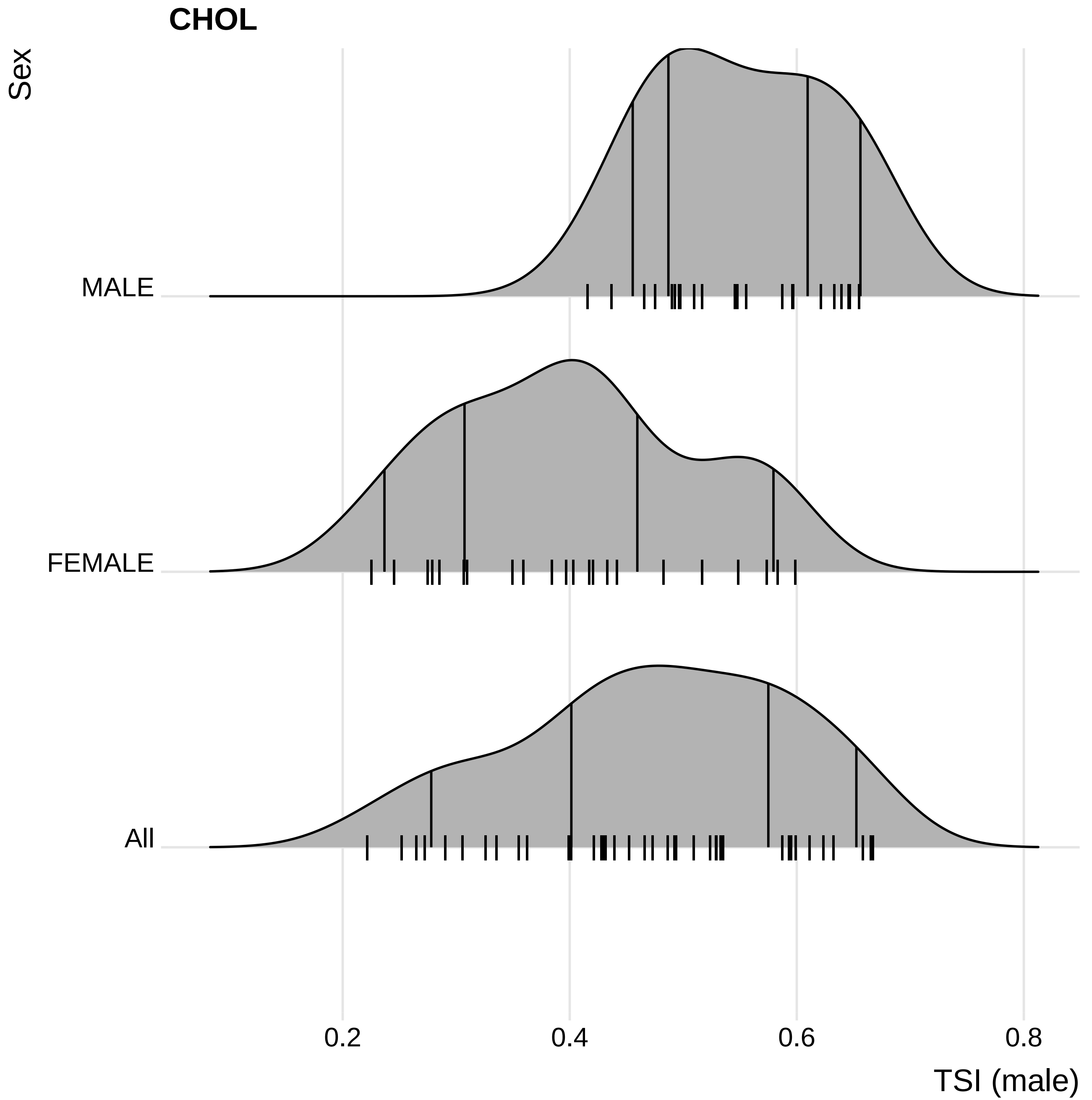
CHOL
Sex
MALE
FEMALE
All
0.2
0.4
0.6
0.8
TSI (male)

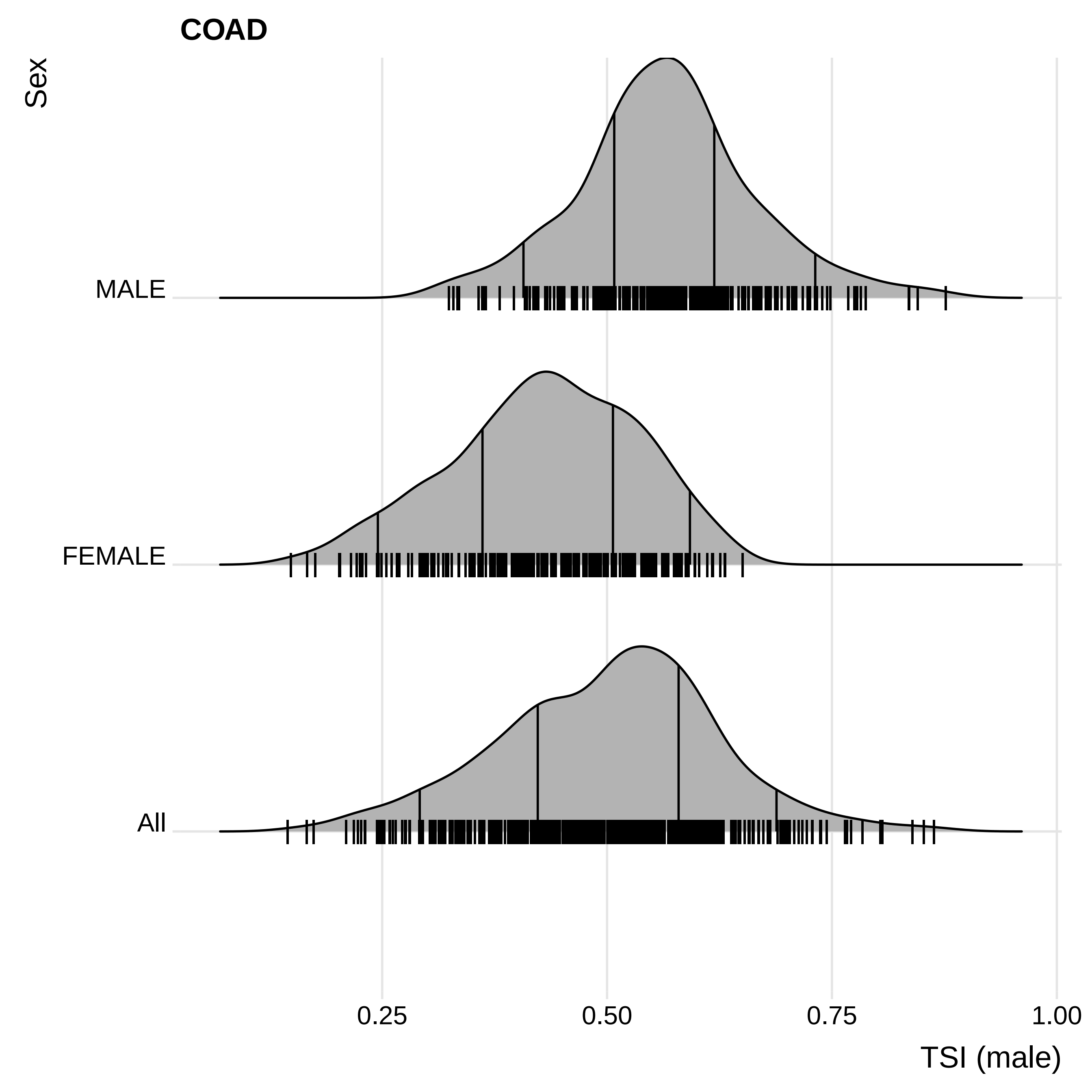

COAD
Sex
MALE
FEMALE
All
0.25
0.50
0.75
1.00
TSI (male)

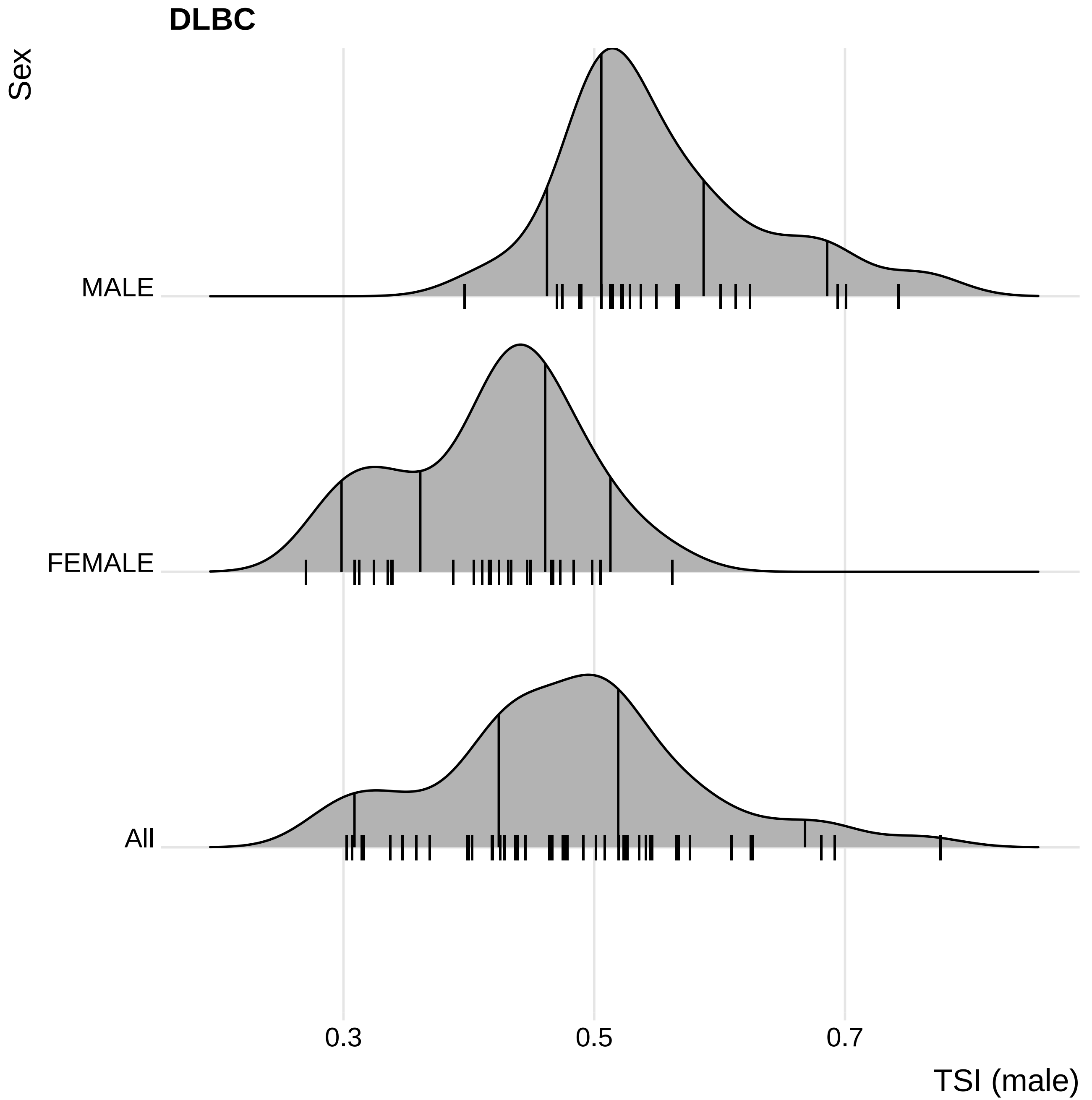
DLBC
Sex
MALE
FEMALE
All
0.3
0.5
0.7
TSI (male)

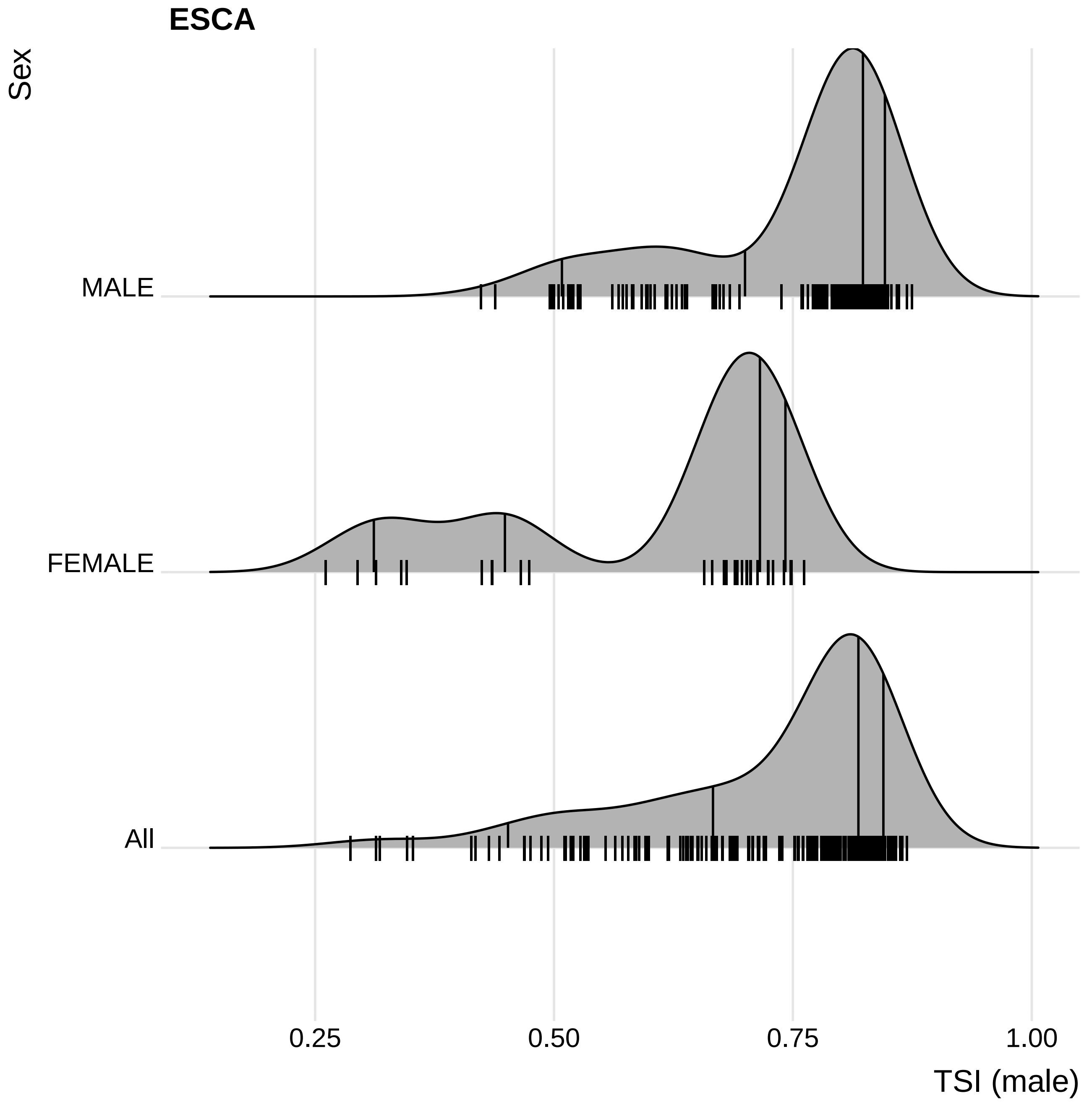

ESCA
Sex
MALE
FEMALE
All
0.25
0.50
0.75
1.00
TSI (male)

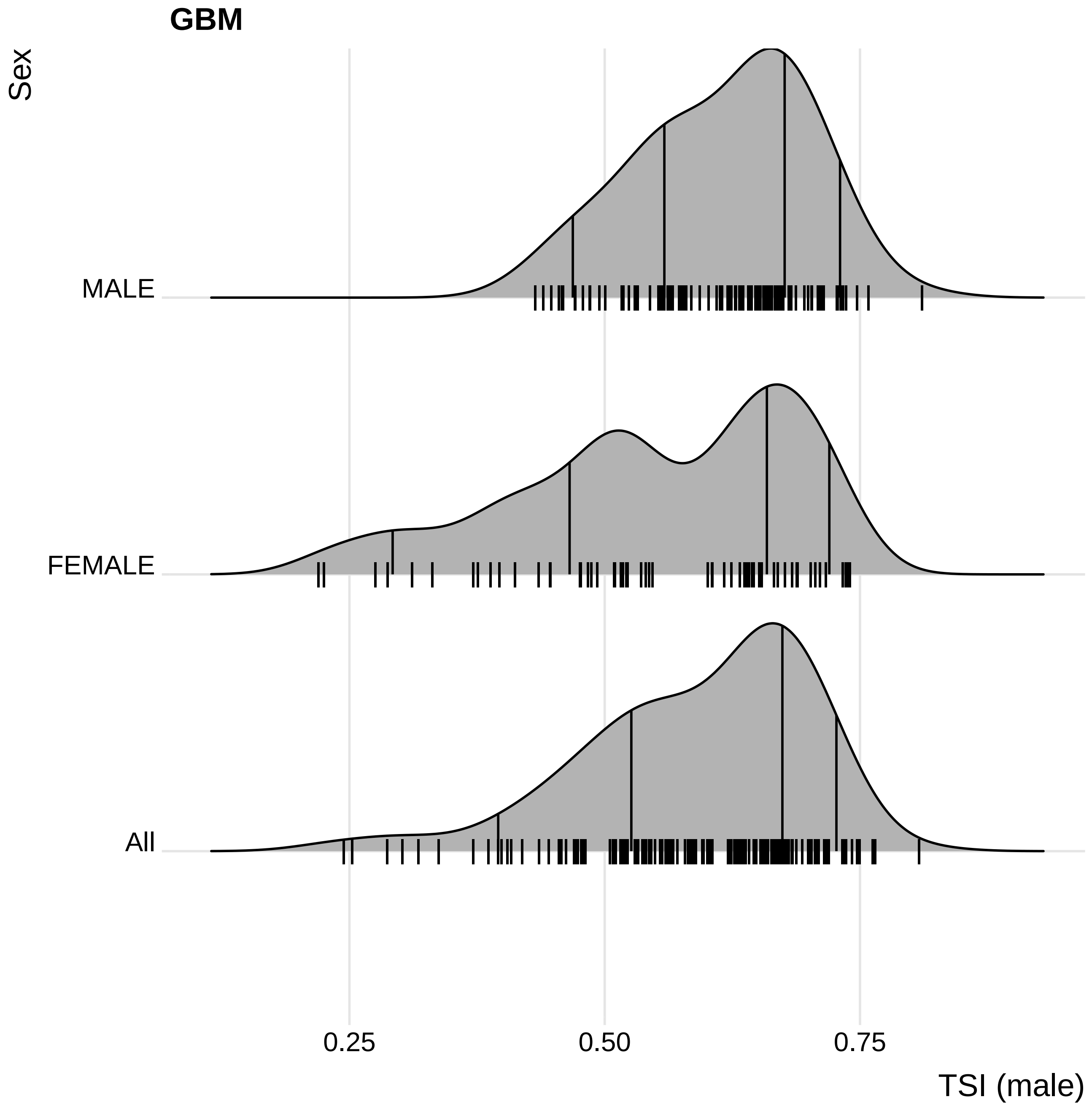
GBM
Sex
MALE
FEMALE
All
0.25
0.50
0.75
TSI (male)

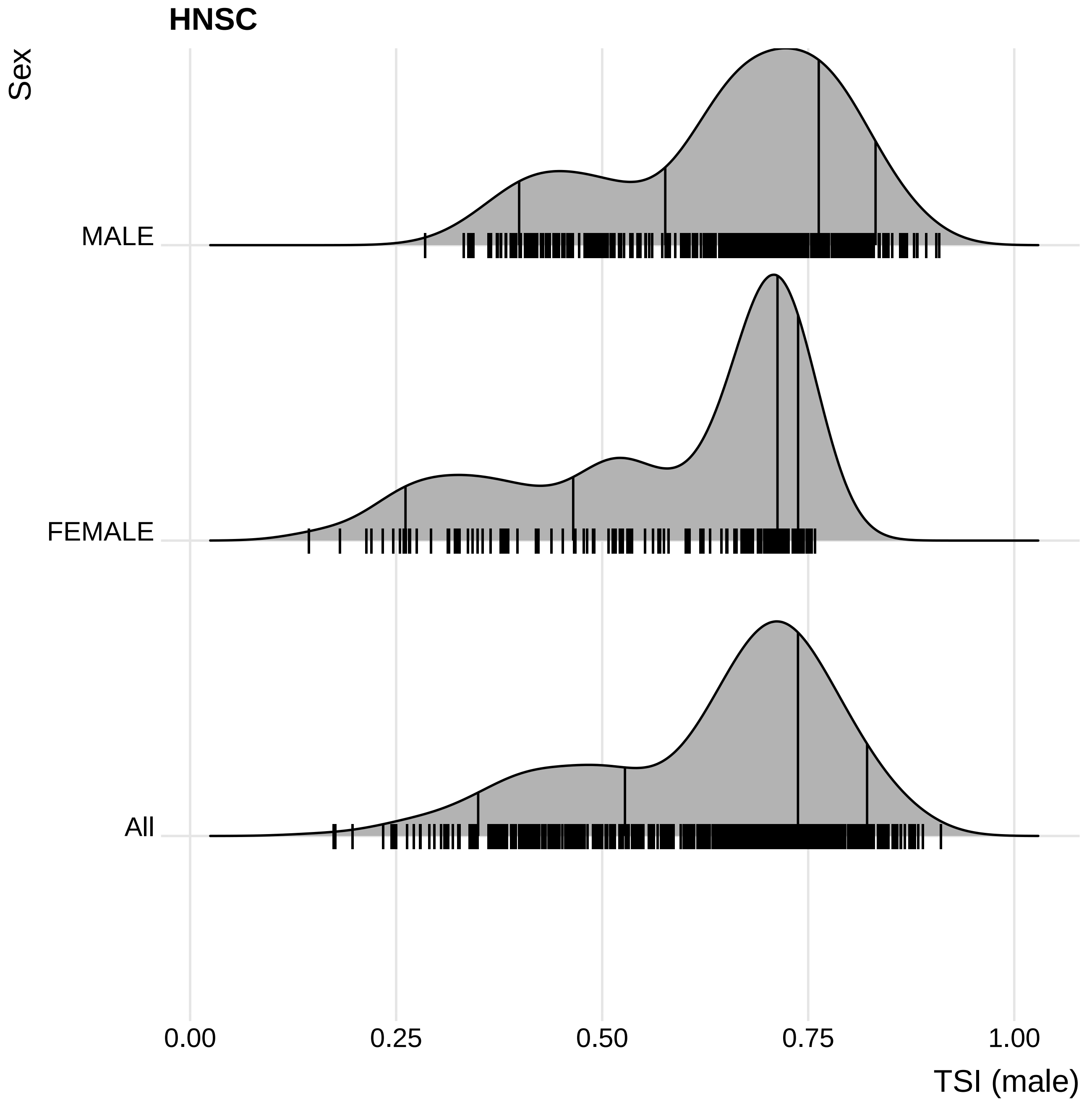
HNSC
Sex
MALE
FEMALE
All
0.00
0.25
0.50
0.75
1.00
TSI (male)

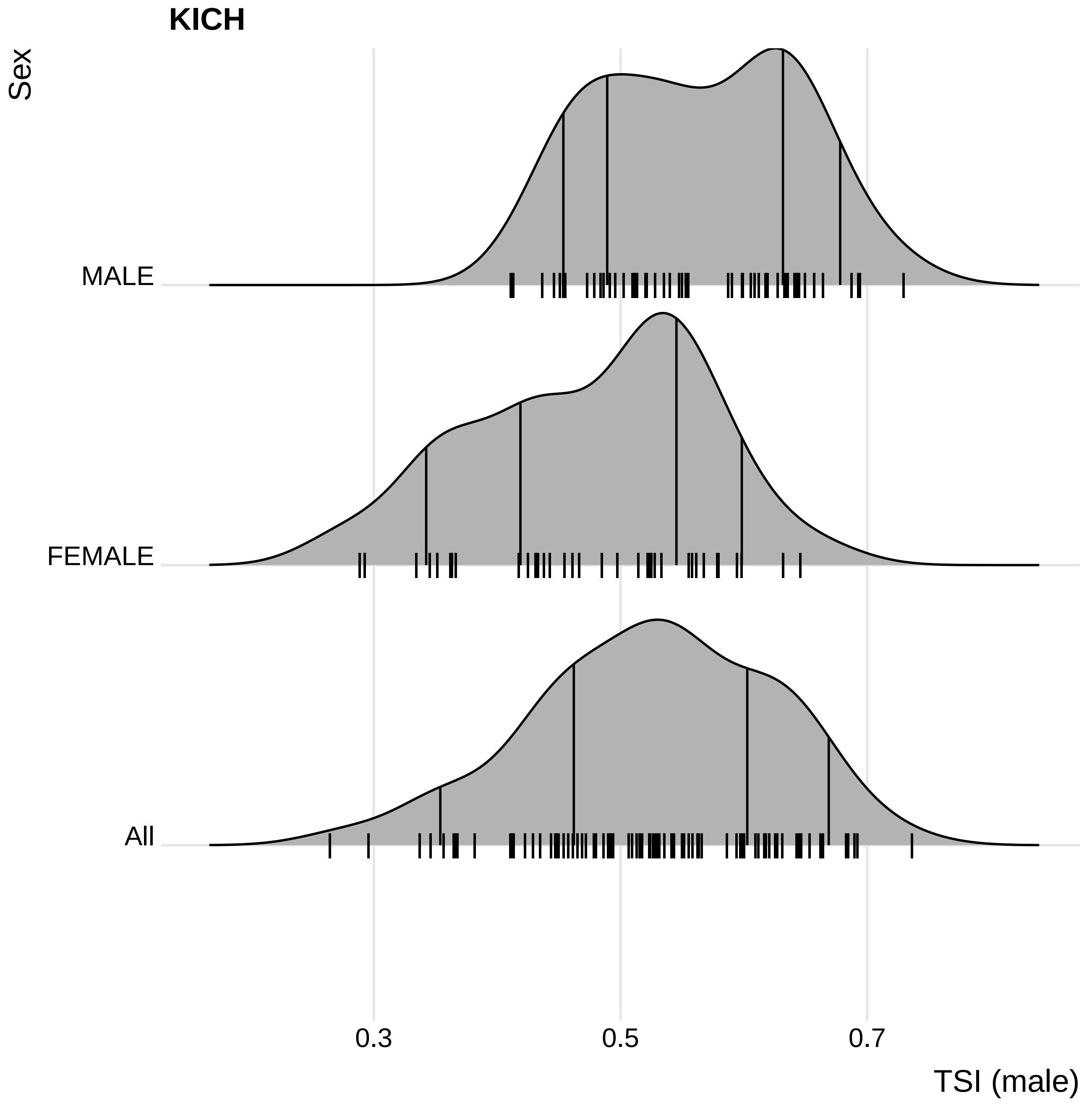

KICH
Sex
MALE
FEMALE
All
0.3
0.5
0.7
TSI (male)

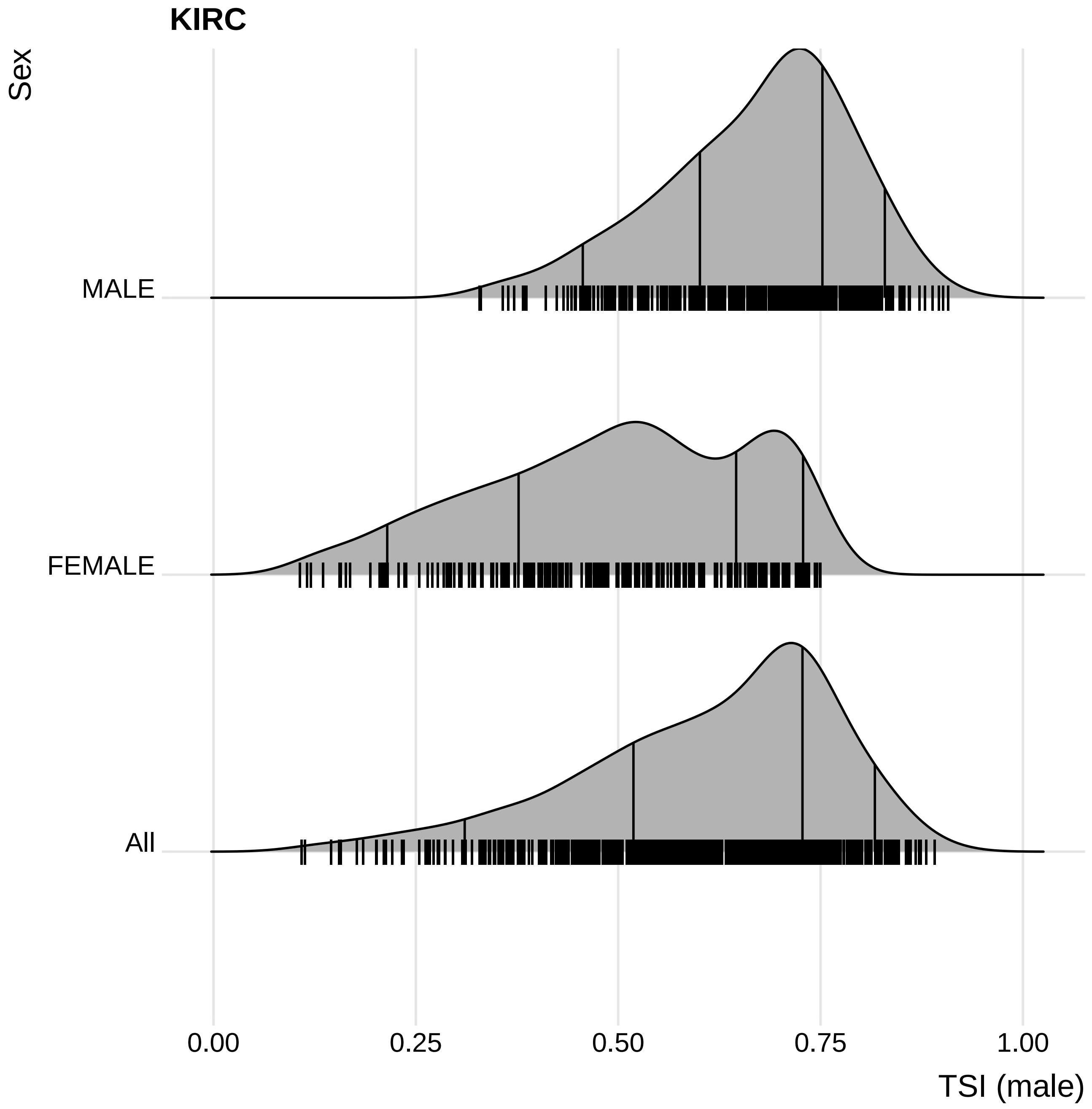

KIRC
Sex
MALE
FEMALE
All
0.00
0.25
0.50
0.75
1.00
TSI (male)

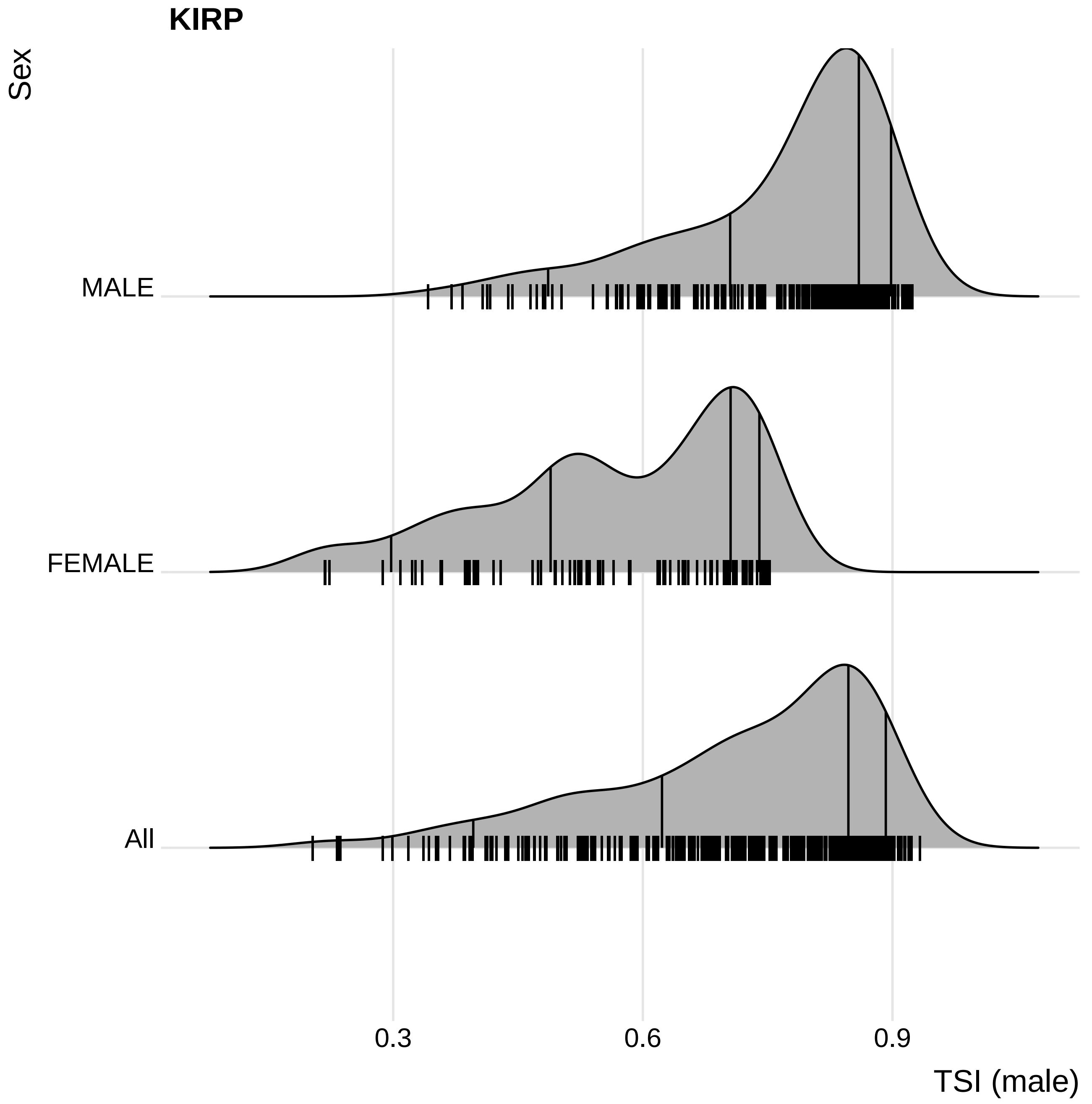
KIRP
Sex
MALE
FEMALE
All
0.3
0.6
0.9
TSI (male)

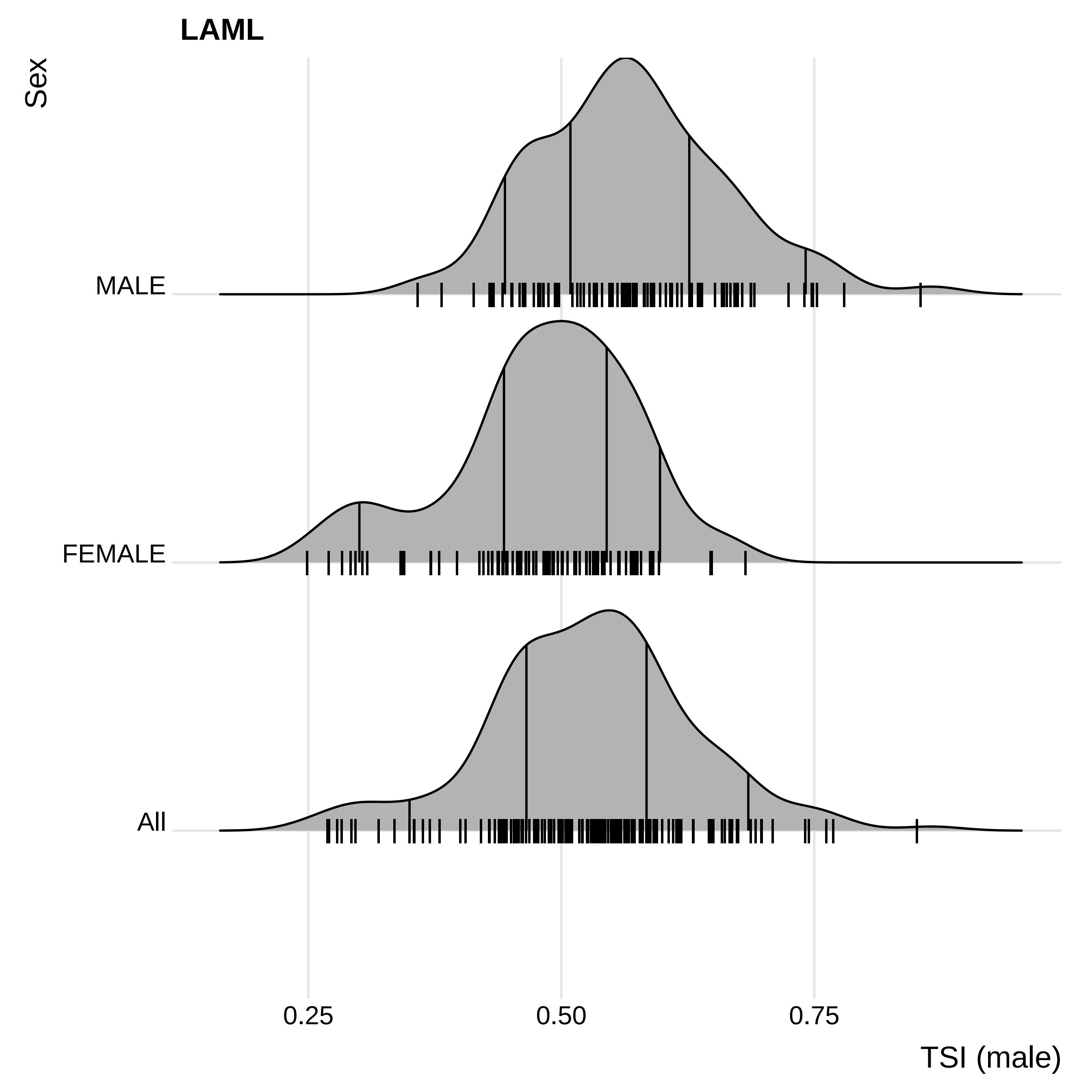
LAML
Sex
MALE
FEMALE
All
0.25
0.50
0.75
TSI (male)

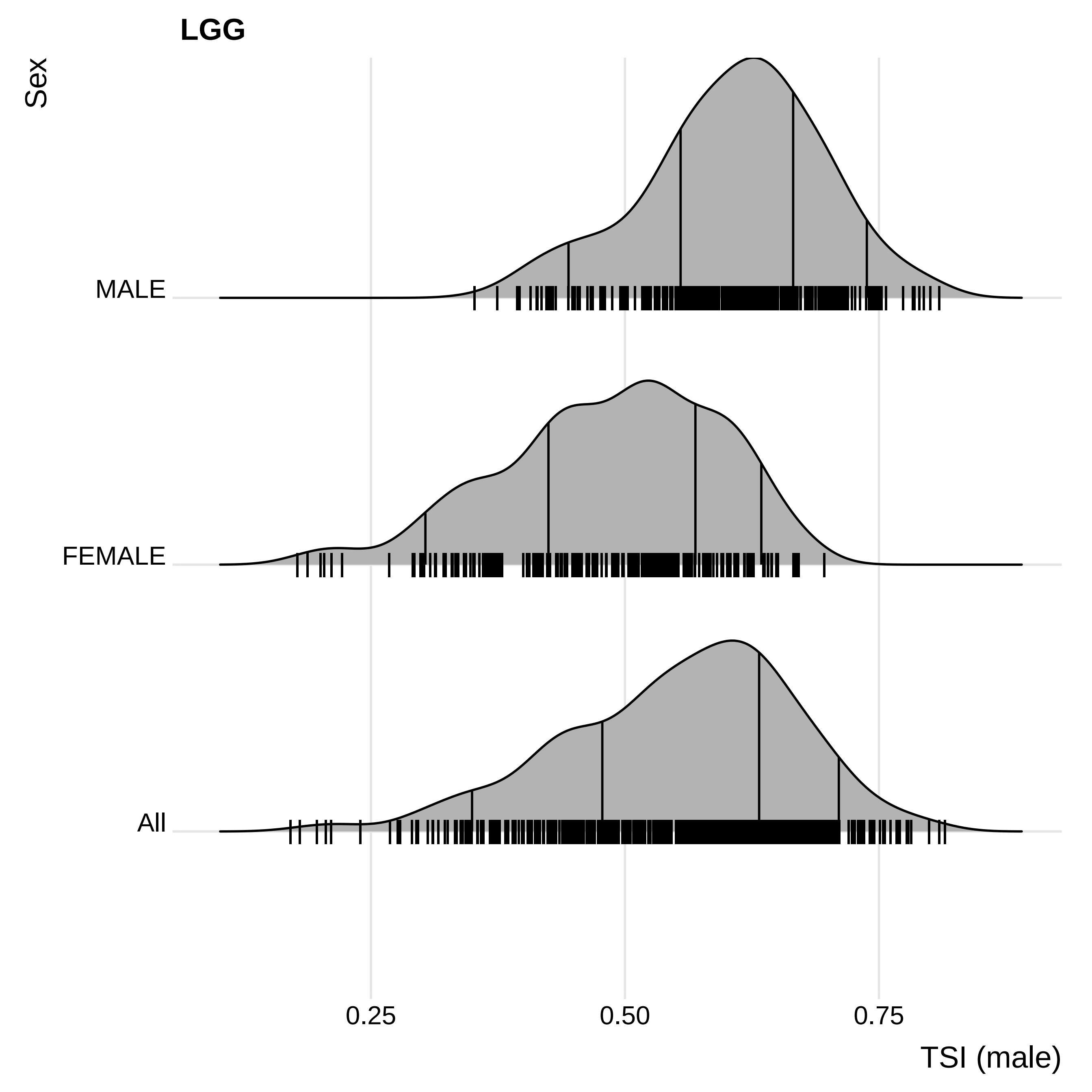
LGG
Sex
MALE
FEMALE
All
0.25
0.50
0.75
TSI (male)

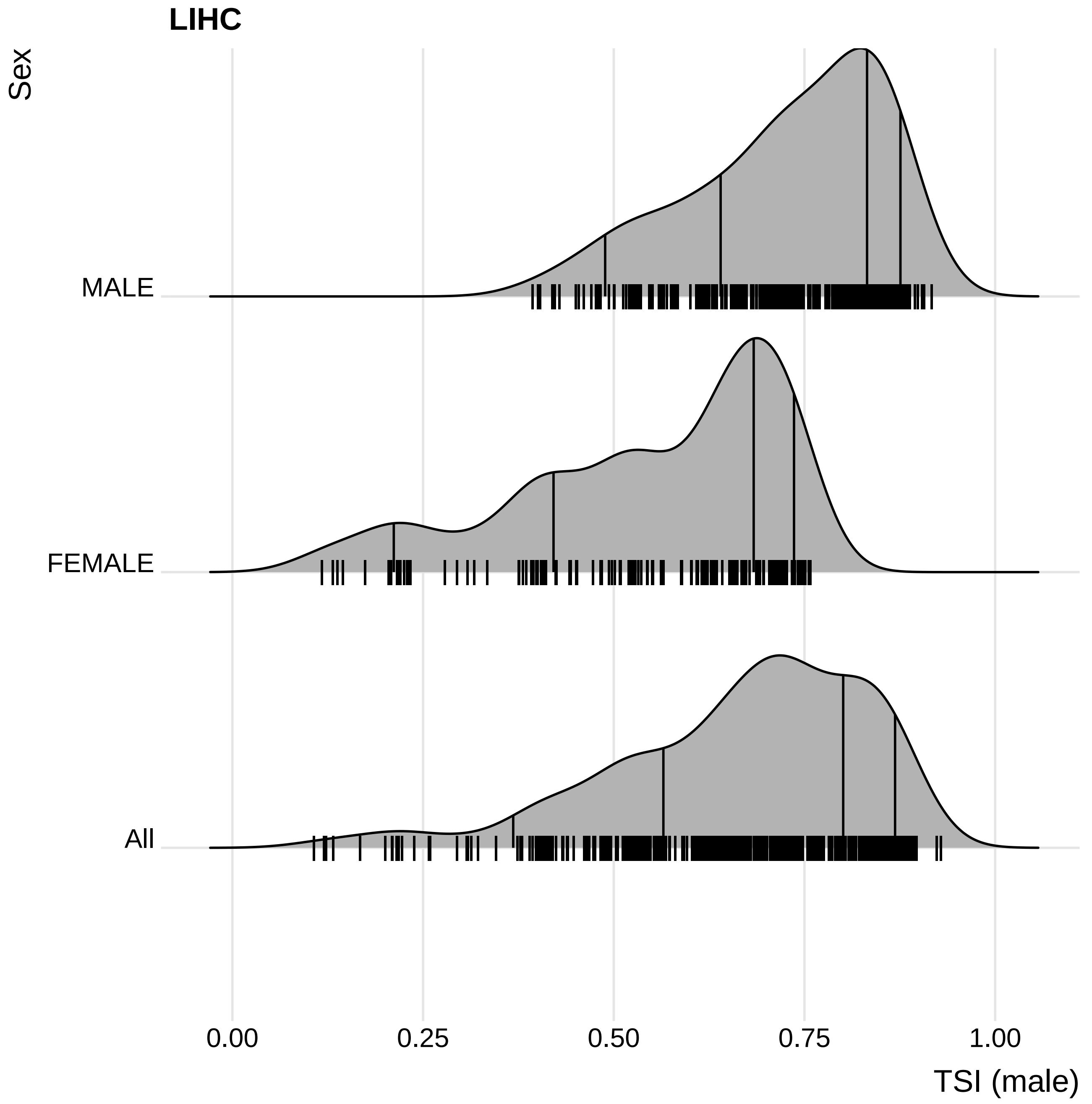

LIHC
Sex
MALE
FEMALE
All
0.00
0.25
0.50
0.75
1.00
TSI (male)

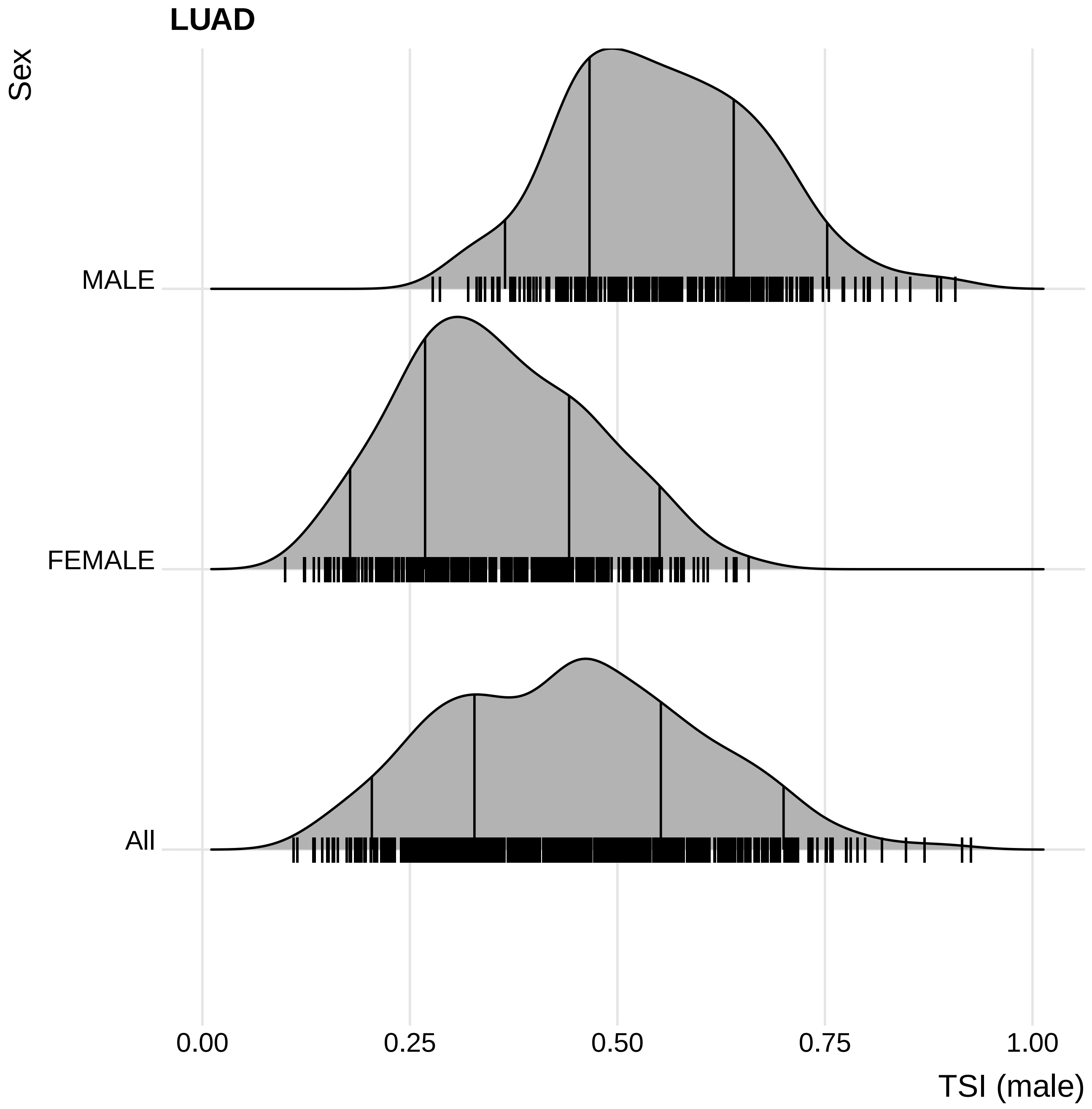

LUAD
Sex
MALE
FEMALE
All
0.00
0.25
0.50
0.75
1.00
TSI (male)

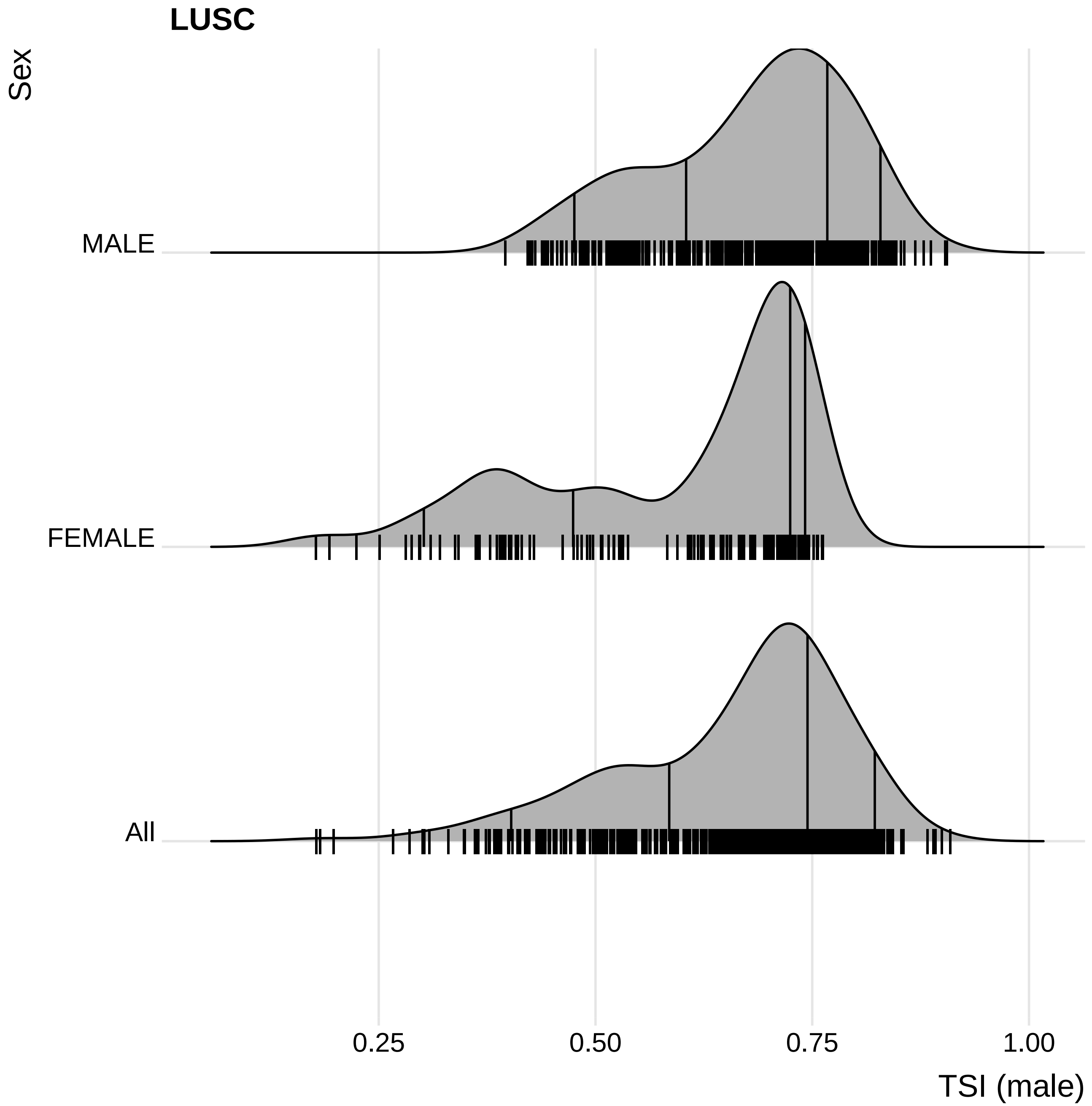

LUSC
Sex
MALE
FEMALE
All
0.25
0.50
0.75
1.00
TSI (male)

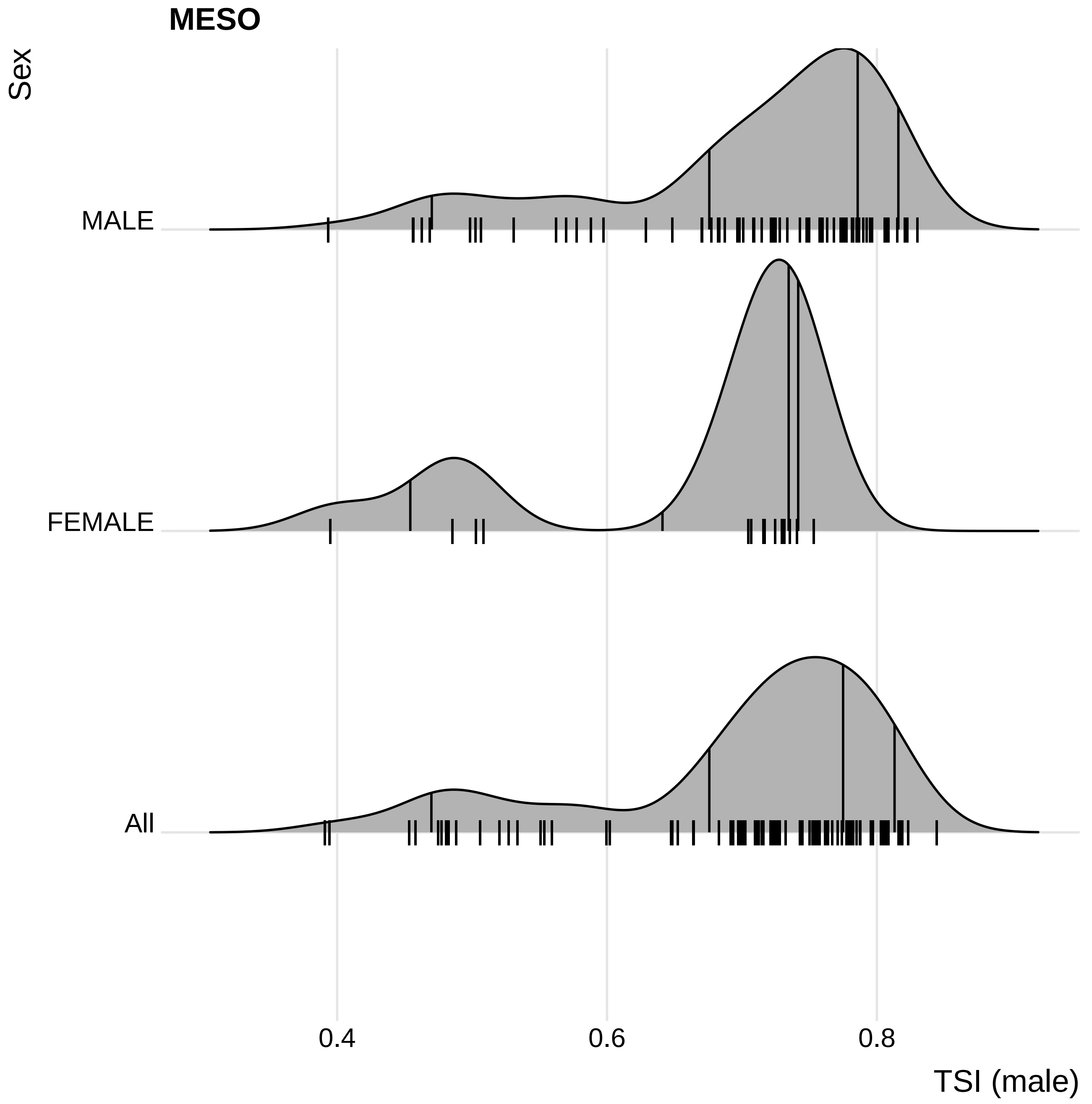
MESO
Sex
MALE
FEMALE
All
0.4
0.6
0.8
TSI (male)

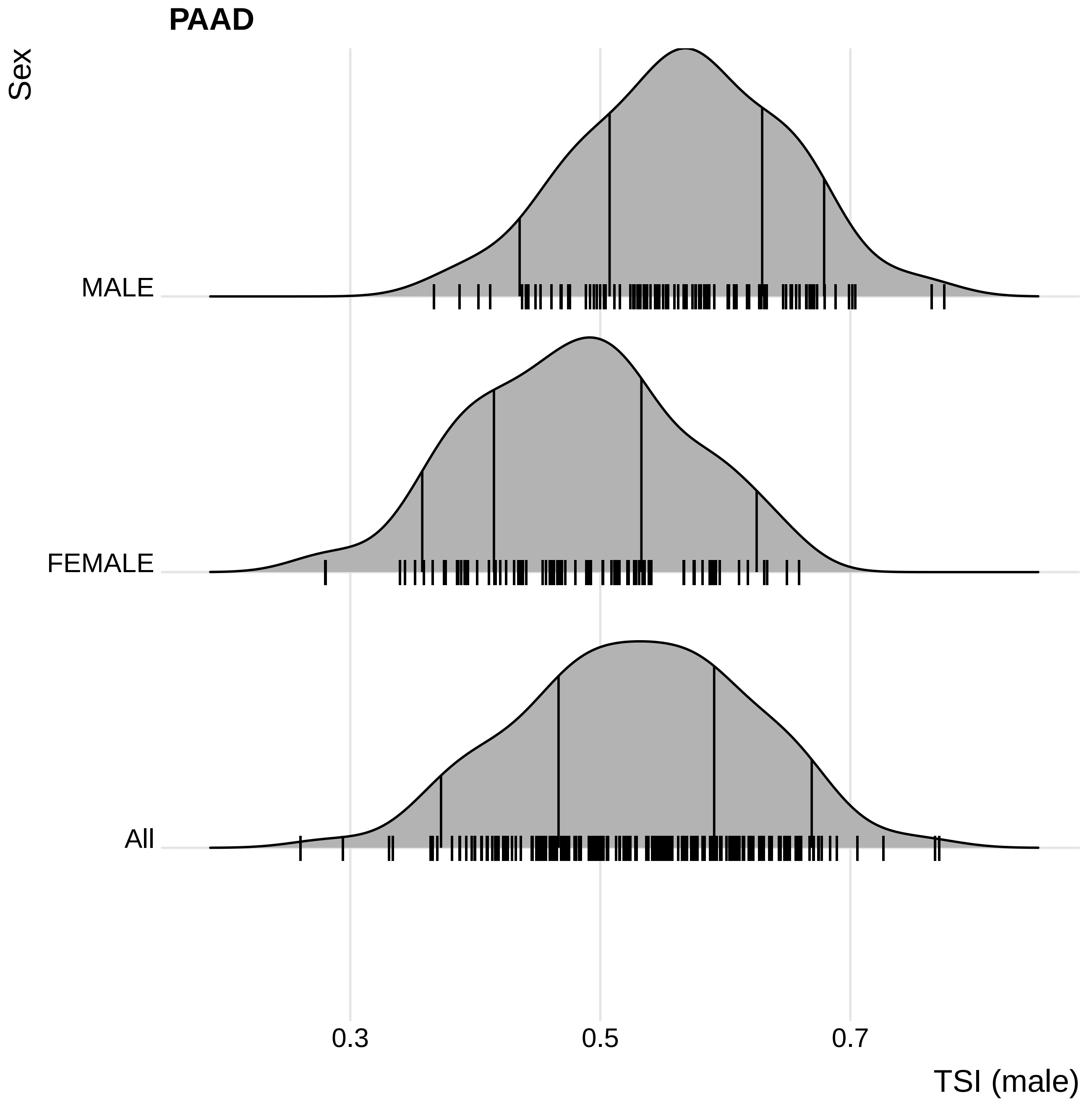

PAAD
Sex
MALE
FEMALE
All
0.3
0.5
0.7
TSI (male)

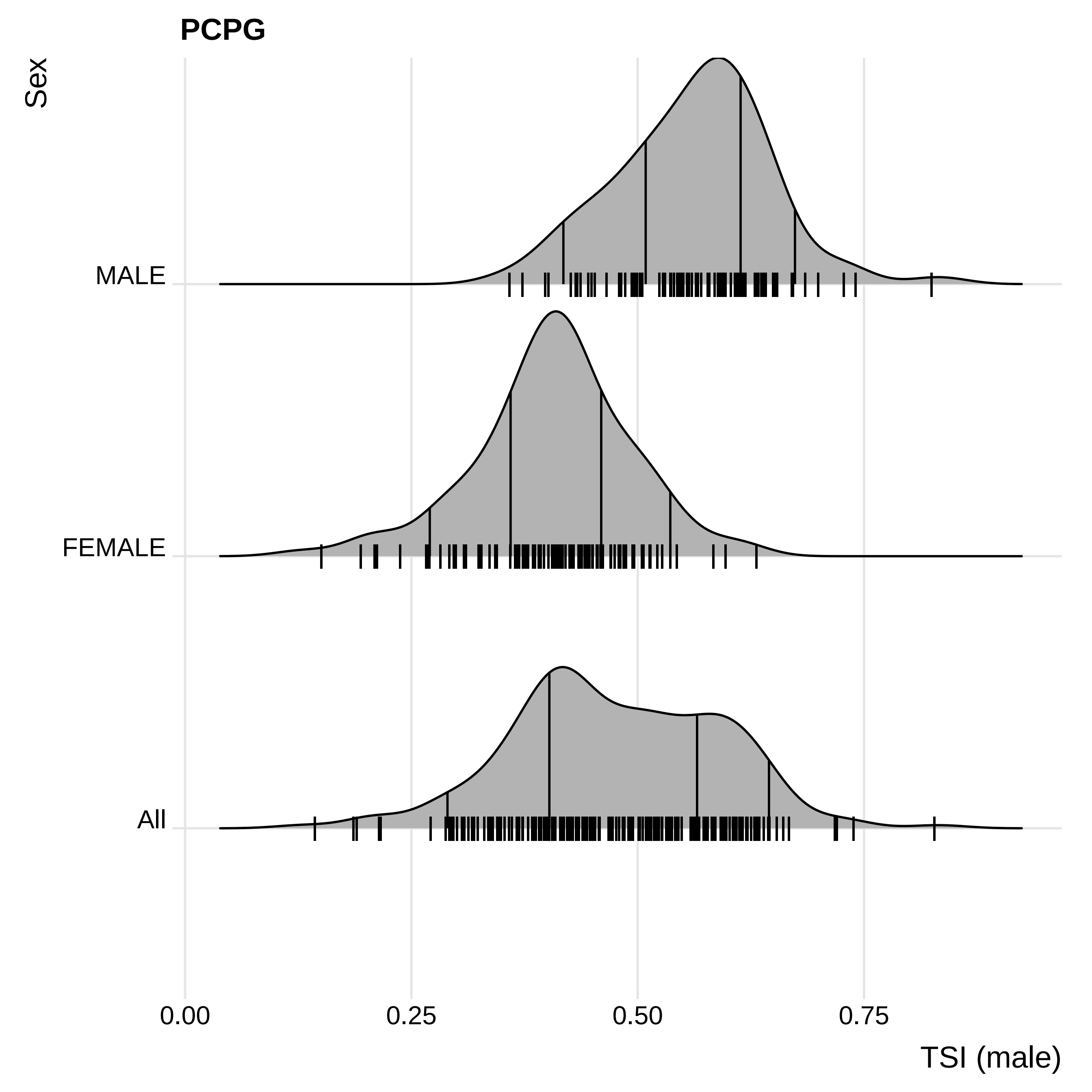

PCPG
Sex
MALE
FEMALE
All
0.00
0.25
0.50
0.75
TSI (male)

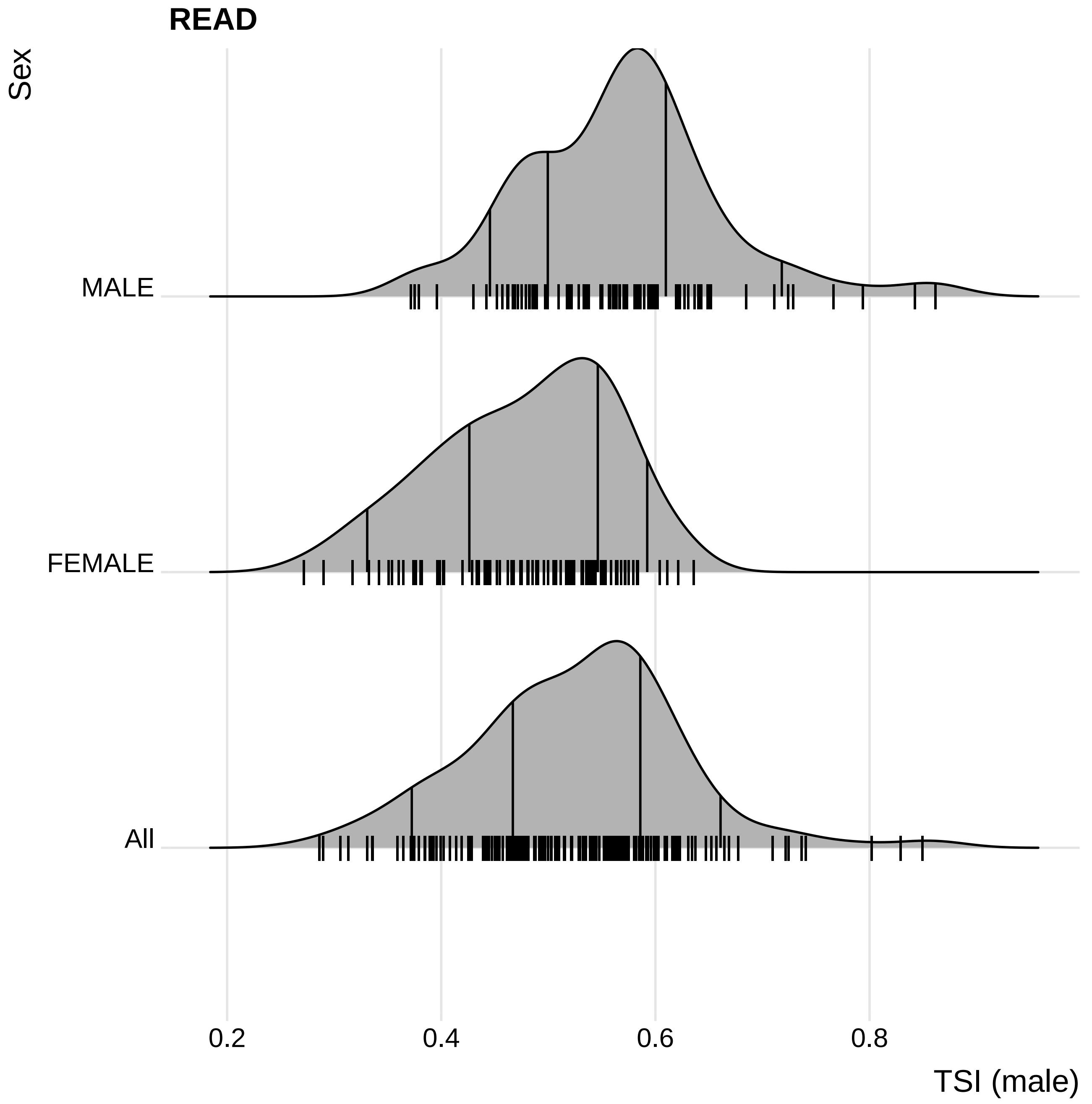
READ
Sex
MALE
FEMALE
All
0.2
0.4
0.6
0.8
TSI (male)

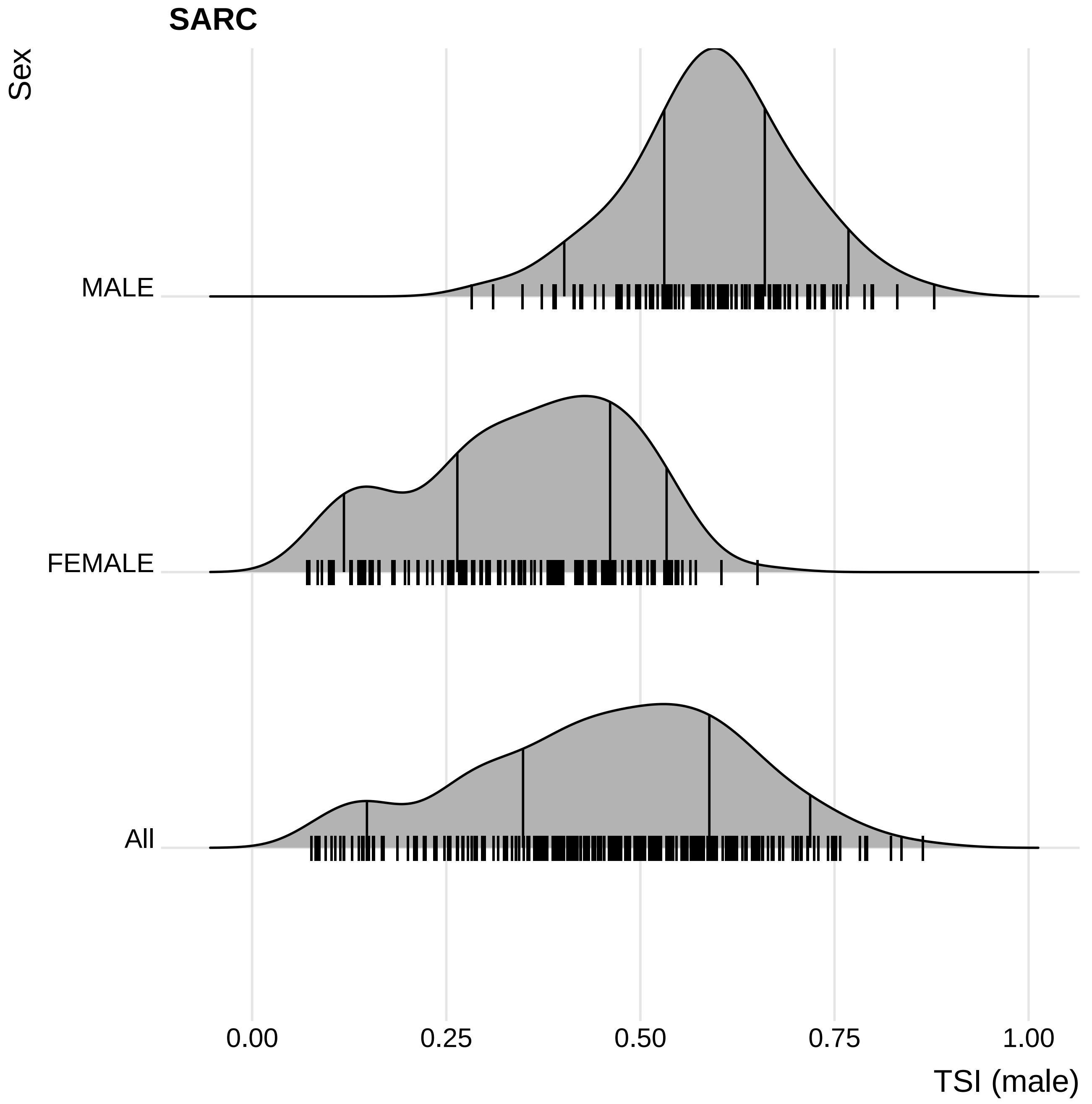
SARC
Sex
MALE
FEMALE
All
0.00
0.25
0.50
0.75
1.00
TSI (male)

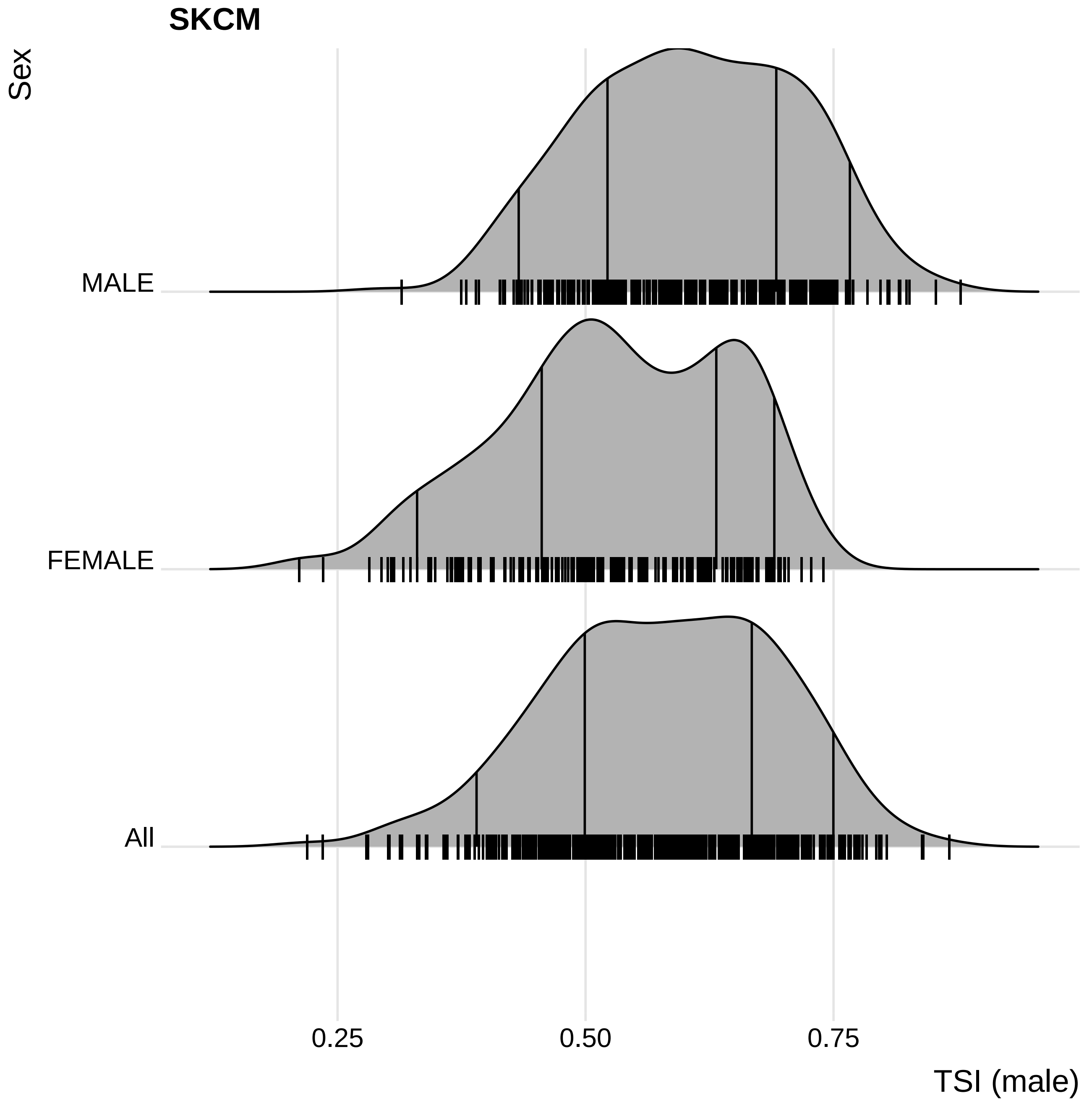
SKCM
Sex
MALE
FEMALE
All
0.25
0.50
0.75
TSI (male)

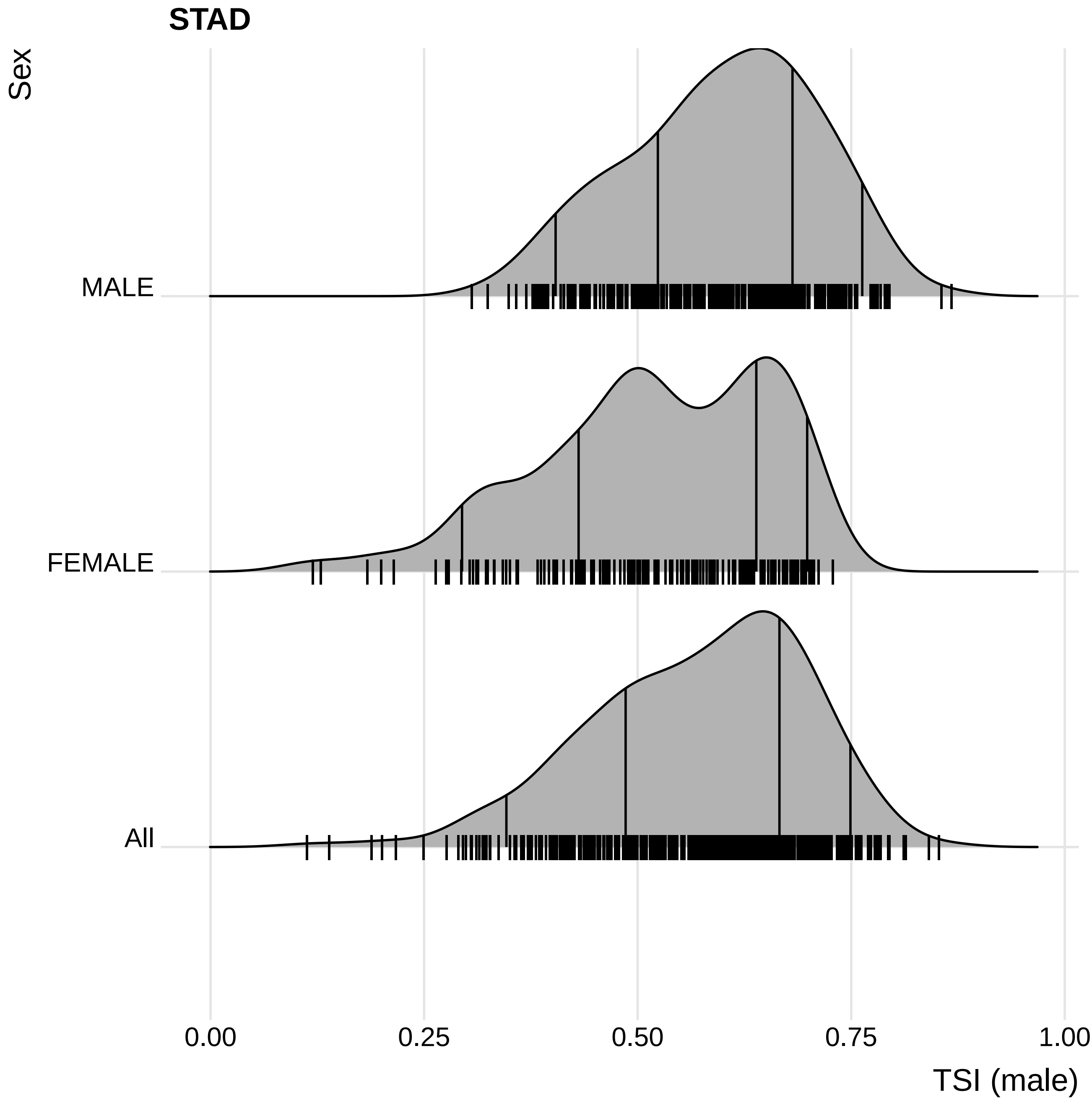

STAD
Sex
MALE
FEMALE
All
0.00
0.25
0.50
0.75
1.00
TSI (male)

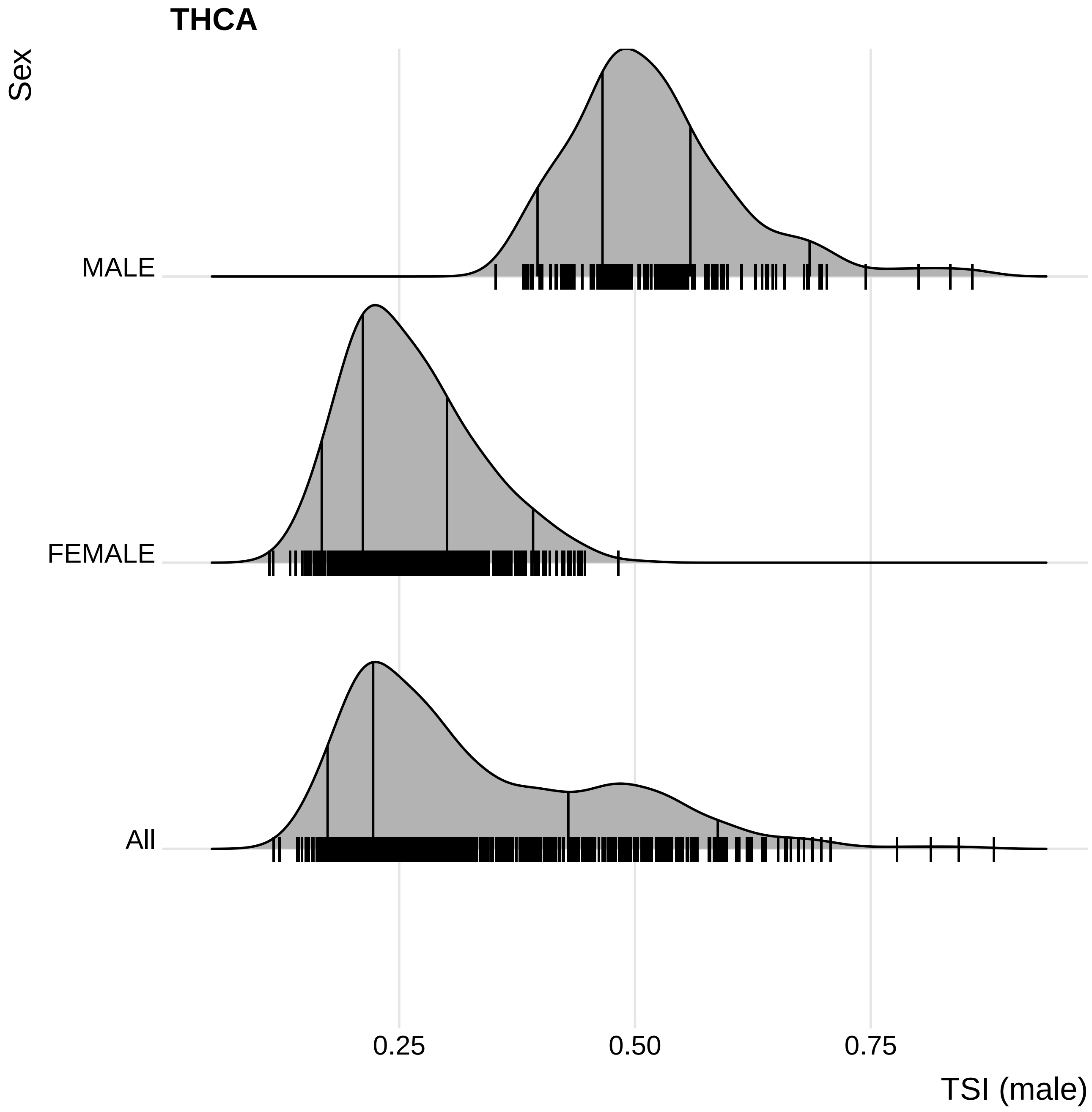
THCA
Sex
MALE
FEMALE
All
0.25
0.50
0.75
TSI (male)

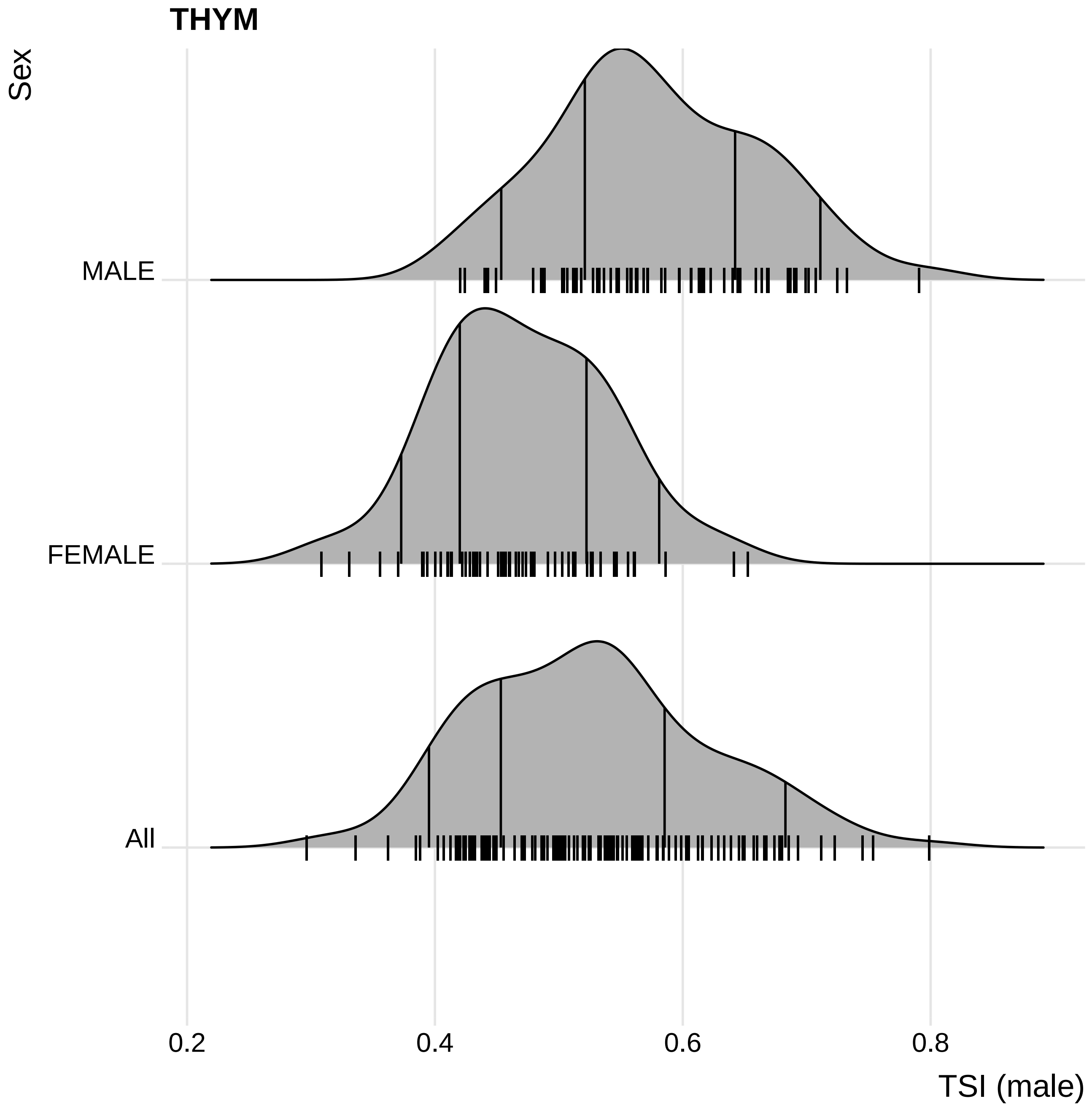

THYM
Sex
MALE
FEMALE
All
0.2
0.4
0.6
0.8
TSI (male)

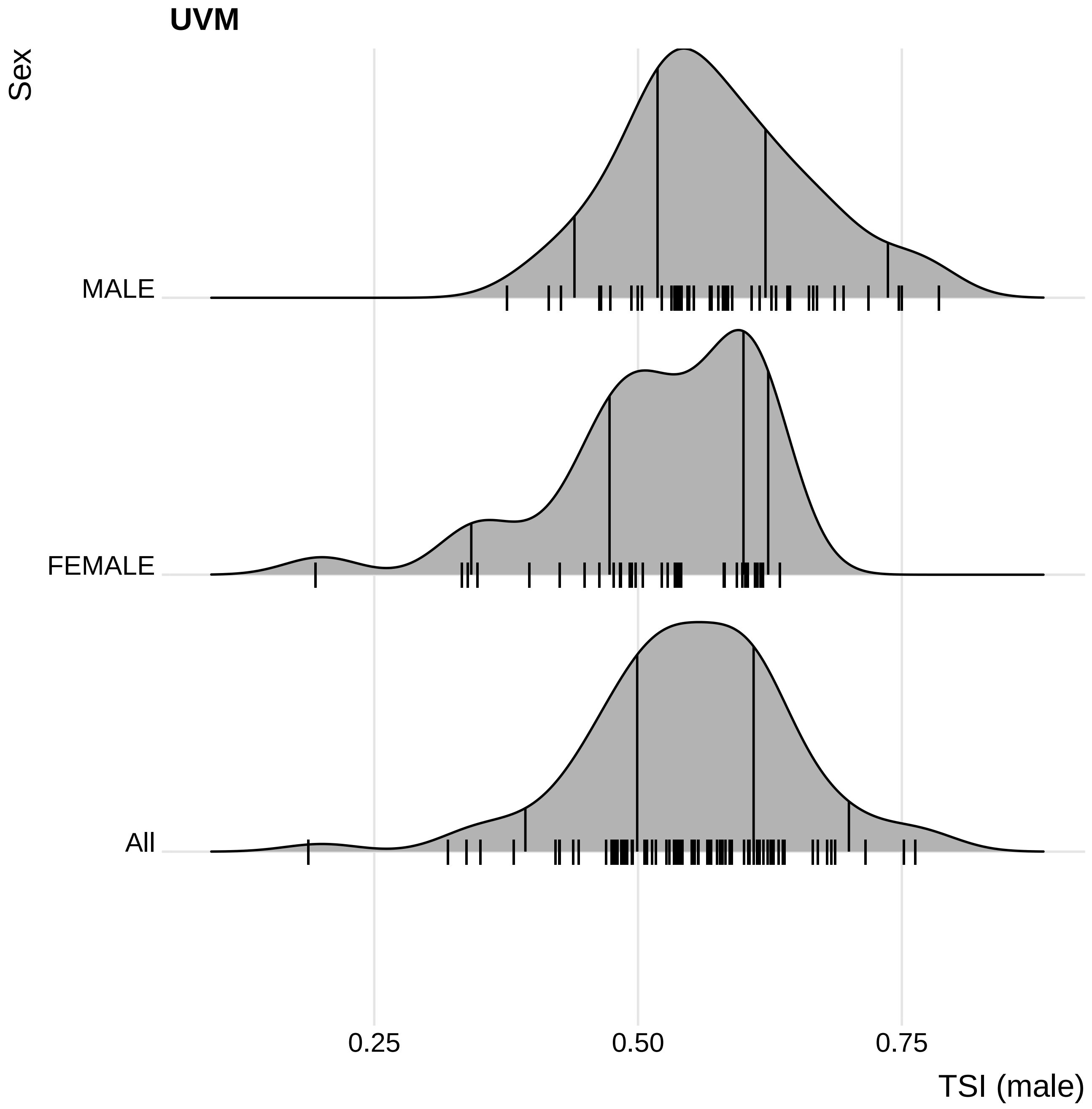
UVM
Sex
MALE
FEMALE
All
0.25
0.50
0.75
TSI (male)

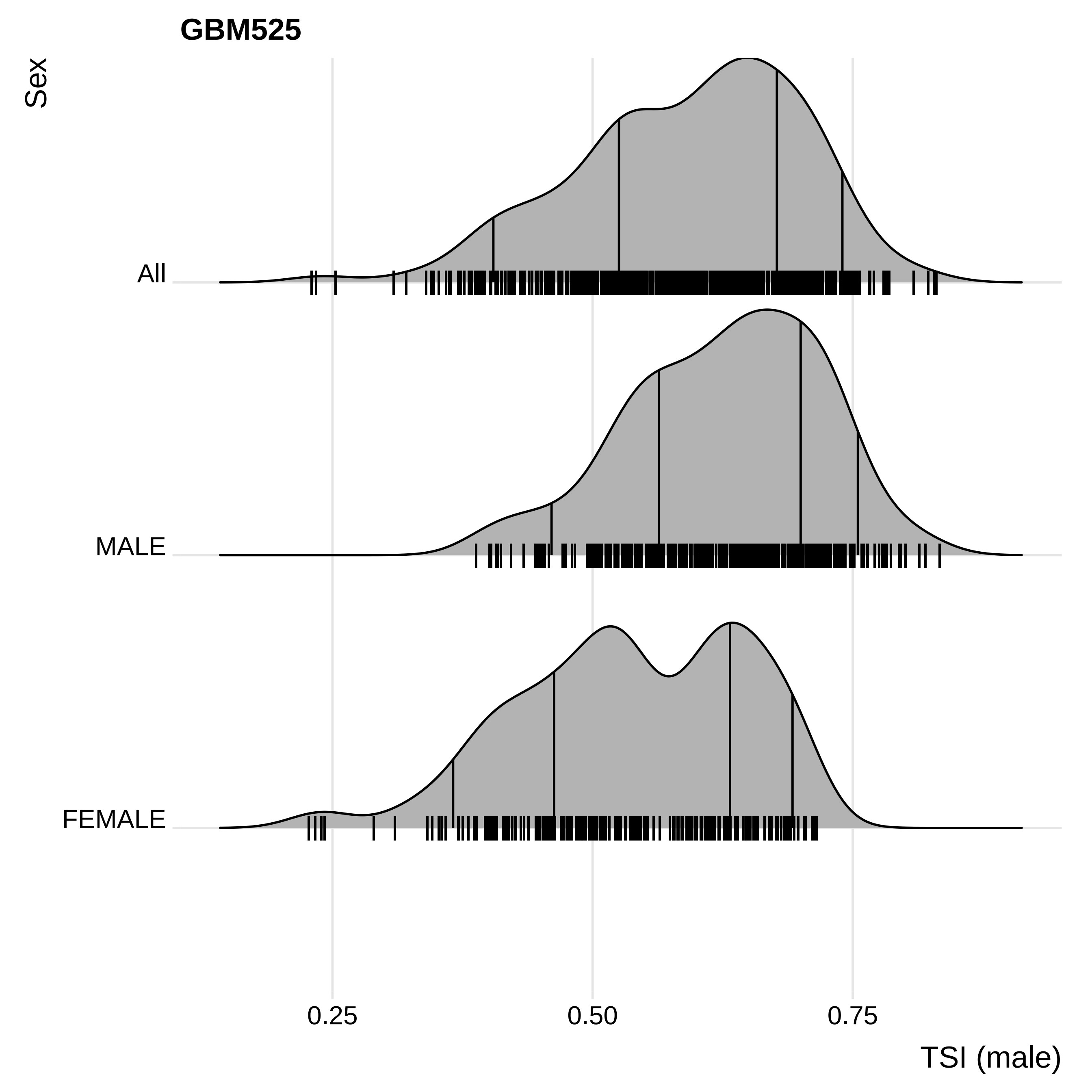
GBM525
Sex
All
MALE
FEMALE
0.25
0.50
0.75
TSI (male)
